# Unveiling prognostic genes and regulatory mechanisms of exosome in prostate cancer: an integrated analysis of bulk transcriptomics and single-cell RNA sequencing data

**DOI:** 10.64898/2025.12.23.25342923

**Authors:** Chunlin Pu

**Author notes:** **Correspondence** Chunlin Pu.

## Abstract

**Objective:** Prostate cancer (PCa) constitutes a considerable public health concern worldwide, primarily attributable to its elevated mortality rates. Changes in exosome are shown to significantly influence tumor development. This study aimed to investigate the prognostic value of exosome-related genes (ERGs) in PCa.

**Methods:** PCa single-cell RNA sequencing (scRNA-seq) and transcriptome datasets were obtained from public databases, with ERGs extracted from existing literature. Candidate genes were identified by overlapping 6,004 PCa-related differentially expressed genes (DEGs) and 121 ERGs. Multiple algorithms screened prognostic genes to construct and validate a risk model. Function enrichment, immune infiltration, and drug sensitivity analyses were performed for high/low-risk groups, while scRNA-seq determined cell types via prognostic genes.

**Results:** A sum of 36 candidate genes was discovered at the intersection of 6,004 DEGs and 121 ERGs. NOC2L, RPS10, POSTN, and BIRC5 were selected as the prognostic genes. The survival status of PCa patients was effectively predicted by a risk model. The majority of pathways identified as significantly enriched between the 2 groups were related to cellular functions. Additionally, 7 differential immune cell types were identified between the 2 groups. RPS10 demonstrated the most significant negative correlation with immature dendritic cells. Chemotherapy drugs were more effective for PCa patients classified as low-risk group. Finally, epithelial cells, endothelial cells, and T cells were considered as key cells and played a critical role in PCa.

**Conclusion:** NOC2L, RPS10, POSTN, and BIRC5 were identified associated with exosome in PCa, providing a strong reference for exosome mechanisms in PCa.

## 1. Introduction

Prostate cancer (PCa) is the second most common malignant tumor in men globally, with rising incidence linked to population aging and dietary changes[1, 2]. In the United States, prostate cancer incidence rose by 3% annually from 2014 to 2019, with projections for 2023 indicating 288,300 new cases and 34,700 deaths[3]. In China, prostate cancer (PCa) constitutes 14. 1% of new male cancer cases and is the fifth leading cause of cancer-related deaths, responsible for 6. 8% of male cancer fatalities [4]. This disease exhibits complex pathological features, and its multifocal growth pattern poses challenges to early diagnosis. In terms of treatment, early-stage PCa is mainly managed through radical prostatectomy, while advanced PCa is managed with androgen deprivation therapy (ADT), radiotherapy, and chemotherapy [5]. Therapeutic resistance is unavoidable, and its emergence in advanced prostate cancer, including castration-resistant prostate cancer (CRPC), poses a significant clinical challenge [5, 6]. This transformation, coupled with the inherent complexity of the disease, underscores the urgency of conducting in-depth research into the pathogenesis of PCa, as well as developing novel prognostic biomarkers and targeted therapeutic strategies—an important and pressing task in current oncology research.

Exosomes are lipid bilayer vesicles (30–150 nm) known for their stable membranes and ability to traverse biological barriers, such as the blood-brain barrier, without eliciting immune responses due to their unique lipid composition. They serve as intercellular communicators by transporting bioactive molecules like proteins and RNAs to influence recipient cell functions. Detecting exosomes offers a non-invasive liquid biopsy method for monitoring disease status and progression. Owing to their excellent stability and biocompatibility, exosomes also act as promising drug delivery vehicles [7] and exhibit broad prospects in clinical applications, including gene therapy and immune regulation[8]. The role of exosomes in prostate cancer (PCa) has attracted increasing attention, particularly their regulatory effects on in immune suppression, angiogenesis, metastasis, and therapeutic resistance. As carriers of biomolecules such as nucleic acids and proteins, exosomes can transmit information between cells, thereby influencing cancer progression and treatment outcomes [9, 10]. Exosomes facilitate cancer cell invasion and metastasis by transporting specific miRNAs and proteins. Exosomes released by prostate cancer-associated fibroblasts (CAFs) enhance prostate cancer metastasis through the miR-500a-3p/FBXW7/HSF1 pathway in hypoxic environments[11]. In particular, the long non-coding RNA HOXD-AS1 found in exosomes from prostate cancer aids in the distant spread of the disease through the miR-361-5p/FOXM1 pathway. Exosomes significantly contribute to therapeutic resistance in prostate cancer (PCa). Research indicates that the circular RNA circ-XIAP in prostate cancer exosomes facilitates docetaxel resistance by modulating the miR-1182/TPD52 axis [12]. miRNAs in exosomes are also closely related to chemotherapy resistance; specifically, certain miRNAs modulate drug resistance in PCa cells by regulating key genes (e. g., AR, PTEN)[19]. Exosomes hold significant promise as non-invasive biomarkers for diagnosing and treating prostate cancer. Exosomal proteins and miRNAs are potential biomarkers for early diagnosis and monitoring of prostate cancer progression [13, 14]. Prostate-specific antigen (PSA) and other protein levels in PCa exosomes can effectively differentiate prostate cancer patients from those with benign prostatic hyperplasia (BPH)[15]. Exosomal miRNA-1246 is a potential biomarker for aggressive prostate cancer, with its expression level closely linked to the disease’s aggressiveness and prognosis [16].

Single-cell RNA sequencing (scRNA-seq) is a robust transcriptomic technology that allows for an in-depth, unbiased analysis of cells, facilitating the characterization of intricate biological processes at the single-cell level. It uncovers the heterogeneity and evolutionary trajectories of infiltrating immune cells and cancer cells, while dissecting the composition and functions of cellular populations within the tumor microenvironment (TME)[17]. Such high-resolution sequencing not only deepens the understanding of tumorigenesis but also provides new insights for anticancer immunotherapy. Compared with traditional bulk sequencing, scRNA-seq offers unique advantages: revealing intercellular heterogeneity, identifying rare cell populations, and tracking the dynamic changes of cells under different physiological and pathological condition[18]. This technology is a leading approach for analyzing RNA transcript diversity and complexity in single cells, and for identifying cell type composition in complex organisms [19].

This study focuses on exosome-related prognostic genes in PCa. It screens core genes by integrating public databases and employs bioinformatics methods to analyze their exosome-mediated regulatory mechanisms (e. g., involvement in key pathological processes such as immunosuppression, metastasis, or drug resistance). A prognostic risk model utilizing these genes is developed for disease evaluation. Single-cell data characterize gene expression in malignant cells, fibroblasts, and immune cells, elucidating exosomes’ role in intercellular signaling within the tumor microenvironment. Finally, the clinical significance of these genes is verified using clinical samples.

This study offers insights into the molecular mechanisms of exosomes in PCa progression and establishes a theoretical basis for developing precise diagnostic markers and targeted therapies using exosome-related genes, aiding the clinical application of precision medicine for PCa.

## 2. Materials and methods

### 2.1 Data collection

Data on RNA-seq, somatic mutations, clinical characteristics, and survival information of prostate cancer patients were obtained from the The Cancer Genome Atlas (TCGA) database (https://portal.gdc.cancer.gov/) on March 20, 2025. The TCGA-PCa dataset contained 51 normal tissue samples and 484 PCa tissue samples. Among the 484 PCa samples, survival information was available for 408 patients. The 408 samples were randomly split into a training set of 285 samples and a testing set of 123 samples, following a 7:3 ratio. In the Gene Expression Omnibus (GEO) database (https://www.ncbi.nlm.nih.gov/geo/), 248 prostate cancer tissue samples were selected from the GSE116918 dataset on the GPL25318 platform. The 248 PCa samples, complete with survival details and gene expression data, were available for use as a validation set. The scRNA-seq dataset GSE193337 (Sequencing platform: GPL20301) contained 4 PCa samples and 4 normal samples. Furthermore, 121 ERGs were identified from the published literature [20] (**S1 Table**).

### 2.2 Analysis of differential expression

Differential expression analysis to identify Differentially Expressed Genes (DEGs) in the TCGA-PCa dataset was conducted using DESeq2 (v 3. 54. 0) [21] on PCa and normal samples, with criteria of |log2 FC| > 0. 5 and P. adjust < 0. 05. The ‘ggplot2’ package (v 3. 5. 1) [22] was used to create a volcano plot of DEGs, highlighting the top 10 genes with the most significant up- and down-regulation based on |log2 FC|. Subsequently, a heatmap of these genes was generated using the ‘pheatmap’ package (v 0. 7. 7)[23].

### 2.3 Identification, functional enrichment, PPI network of candidate genes

The ‘ggvenn’ package (v 0. 1. 10) was used to identify candidate genes by intersecting DEGs with estrogen-related genes (ERGs) in PCa. Subsequently, to explore the biological pathways of these genes, candidate genes were analyzed by gene ontology (GO) and Kyoto Encyclopedia of Genes and Genomes (KEGG) using the “clusterProfiler” (v 4. 10. 1) [24] (P < 0. 05). A protein-protein interaction (PPI) network was constructed using the STRING database (https://string-db.org/) with candidate genes as inputs and an interaction score threshold exceeding 0. 15. This approach aimed to elucidate the interactions among genes at the protein level. The identified interactions were then visualized using Cytoscape (v 3. 10. 2) [25].

### 2.4 Identification of prognostic genes

A univariate Cox regression analysis using the ‘survival’ package (v 3. 7. 0) [26] was conducted on candidate genes in the training set to identify those significantly correlated with overall survival (OS) in PCa, with criteria of hazard ratio (HR) ≠ 1 and P < 0. 05. For the visualization of the outcomes, the “forestplot” (v 3. 1. 3) [27] was used. Candidate prognostic genes were further subjected to the proportional hazards (PH) assumption test. Genes with a P < 0. 05 were filtered out, while those with a P > 0. 05 were retained. The ‘glmnet’ package (v 4. 1. 4) [28] was used for Least Absolute Shrinkage and Selection Operator (LASSO) regression analysis on the remaining genes. Prognostic genes were selected by identifying the optimal lambda value, corresponding to minimal error, through cross-validation.

### 2.5 Construction and validation of a risk model

In the training set, a prognostic genes based risk model was constructed using the following formula:

The coef and expr denote the risk coefficient and gene expression, respectively. The optimal threshold, derived from the risk score in the training set, classified 285 PCa patients into high-risk (HRG) and low-risk (LRG) groups. Kaplan-Meier survival curves were created using ‘survminer’ (v 0. 4. 9) [26] to examine OS rate differences between HRG and LRG, with significance assessed by the log-rank test (P < 0. 05). The “timeROC” package (v 0. 4) [29] was employed to generate ROC curves with an AUC greater than 0. 6 for 1-, 2-, and 3-year intervals. In conclusion, a further evaluation assessed the risk model’s accuracy and generalizability. The model was validated using both the testing and validation sets.

### 2.6 Independent prognostic analysis and construction of the nomogram

A comparative analysis of risk score among 4 subgroups defined by clinical characteristics was conducted within the training dataset utilizing the Wilcoxon test (P < 0. 05). Independent risk factors were identified by analyzing clinical characteristics and risk scores using univariate and multivariate Cox regression (HR ≠ 1, P < 0. 05) and verifying the proportional hazards assumption (P > 0. 05). A nomogram was constructed using the “rms” package (v 7. 0. 0) [29] to predict 1-, 2-, and 3-year PCa survival probabilities based on independent risk factors. To assess the reliability of this nomogram, calibration curves were generated with the utilization of “rms” (v 7. 0. 0).

### 2.7 gene set enrichment analysis (GSEA) and immune microenvironment analysis

GSEA was carried out with the objective of elucidating the biological functions in HRG and LRG of PCa patients. The dataset ‘c2. KEGG. v2022. 1. Hs. symbols’ was chosen from the MSigDB (https://www.gsea-msigdb.org/gsea/msigdb). The training set analysis utilized DEseq2 (v 3. 54. 0) to evaluate differential expression between two groups. GSEA was performed using “clusterProfiler” (v 4. 10. 1) with criteria of |NES| > 1 and P < 0. 05.

According to the ssGSEA algorithm, the “GSVA” (v 1. 50. 0) [30] was utilized to determine the infiltration scores of 28 immune cells [31] between 2 groups in the training set. The Wilcoxon test was used to assess differences in immune-infiltrating cell populations between the two groups (P < 0. 05). The ‘psych’ package (v 2. 4. 3) [32] was employed to assess correlations and significance among various immune cells and between prognostic genes and immune cells, with criteria of |correlation| > 0. 3 and P < 0. 05.

### 2.8 Gene mutation analysis

We employed ‘maftools’ (v 2. 18. 0) [33] to evaluate the top 20 mutated genes in both groups concerning tumor mutation burden (TMB). The Wilcoxon test was subsequently employed to evaluate the differences in TMB score between 2 groups (P < 0. 05). The ‘psych’ package (version 2. 4. 3) was employed to assess the correlation between risk score and TMB score, considering significant associations as those with |r| > 0. 3 and P < 0. 05.

### 2.9 Chemotherapeutic drug sensitivity analysis

A total of 138 chemotherapeutic drugs were sourced from the GDSC database (https://www.cancerrxgene.org/) to study their therapeutic effects on prostate cancer patients. The half maximal drug inhibitory concentration (IC50) of each chemotherapeutic drug was calculated through the “pRRophetic” (v 0. 5) [34] based on the 285 PCa patients. The Wilcoxon test was employed to analyze differences in IC50 values between the two groups, with significance set at P < 0. 05. The correlation between the risk score and the top five chemotherapeutic drugs was examined using the ‘psych’ package (v 2. 4. 3), with significant correlations identified at |r| > 0. 3 and P < 0. 05.

### 2.10 Chromosome localization, subcellular localization and regulatory network analyses of prognostic genes

The location of genes on chromosomes is critical for gene expression and regulation. In this study, the “RCircos” (v 1. 2. 2) [35] was referred to determine the chromosome localization of prognostic genes. The subcellular localizations of these genes were predicted using the GeneCards database (https://www.genecards.org/).

To better understand the role of upstream regulators in prognostic gene regulation, target microRNAs (miRNAs) and their corresponding transcription factors (TFs) were predicted using the miRTarBase and JASPAR databases. Cytoscape (v 3. 10. 2) was used to construct the TF/miRNA-mRNA network.

### 2.11 Treatment of scRNA-seq data

The GSE193337 dataset underwent quality control using Seurat (v5. 1. 0) [36] with criteria: 200 < nFeature_RNA < 6,000, 200 < nCount_RNA < 50,000, and percent. mt < 10%. Then the FindVariableFeatures function was used to extract the top 2,000 HVGs for subsequent analysis. Data scaling was performed using the ScaleData function. PCA was performed on the 2,000 most highly variable genes (HVGs). The JackStraw function results were used to select the top 50 principal components with P-values less than 0. 05. The resolution parameter was set to 0. 2, and unsupervised clustering was performed on all dataset cells using the FindClusters and FindNeighbors functions. Then, the outcomes of the clustering were visualized through the application of the UMAP algorithm. Following dimensionality reduction and clustering, cell types within the clusters were identified using the CellMarker2 database and references [37, 38]. The annotation results were visualized using a UMAP plot, illustrating the expression of marker genes across various cell types.

### 2.12 Selection of key cells

The Wilcoxon test was employed to identify key cells by examining the differential expression of prognostic genes in annotated cells from the GSE193337 dataset (P < 0. 05). Key cells were selected for further analysis based on the differential expression of prognostic genes and their critical role in PCa progression and treatment [38–40]. The functional enrichment of key cells was explored through the pathways function in the “ReactomeGSA” (v 1. 16. 1) [41].

### 2.13 Cell communication and pseudotime analyses

The ‘CellChat’ tool (v 1. 6. 1)[42] was utilized to analyze cell communication and interactions among annotated cells and between key cells and others in the GSE193337 dataset. This analysis illustrated the network of interactions among various cell types. The “CellChat” tool (v 1. 6. 1) was employed to detect ligand-receptor interactions among key and other cells. To clarify the differentiation of essential cell types and the expression patterns of prognostic genes at different differentiation stages, UMAP clustering was used for dimensionality reduction and clustering of key cells (resolution = 0. 2). Then, the “Monocle” (v 2. 30. 1) [43] was used for pseudotime analysis, and the cell differentiation trajectory diagram was drawn.

### 2.14 Statistical analysis

All bioinformatics analyses were carried out with the R software (v 4. 3. 2). To compare between distinct groups, the Wilcoxon test was applied (P < 0. 05).

## 3. Results

### 3.1 The 36 candidate genes were associated with exosome in PCa

Differential expression analysis showed that there were 6,004 DEGs between the 2 groups. Among them, 2,865 genes in the PCa group were identified as upregulated genes, and 3,139 genes were identified as downregulated genes (**Figure 1A-1B**). Moreover, a total of 36 shared genes between DEGs and ERGs were identified and selected as candidate genes (**Figure 1C**). The enrichment analysis of the 36 candidate genes revealed associations with 732 GO terms, including ventricular zone neuroblast division (**Figure 1D**, **S2 Table**). Moreover, the KEGG enrichment analysis revealed that 12 pathways were enriched, for instance calcium signaling pathway (**Figure 1E**). Next, a PPI network was constructed, comprising 138 interactions associated with 34 candidate genes, while 2 isolated targets were excluded (**Figure 1F**). Within this network, ITGB1, FGF9, CTSB, and CYP19A1 frequently interacted with other genes at the protein level.

**Figure 1.**
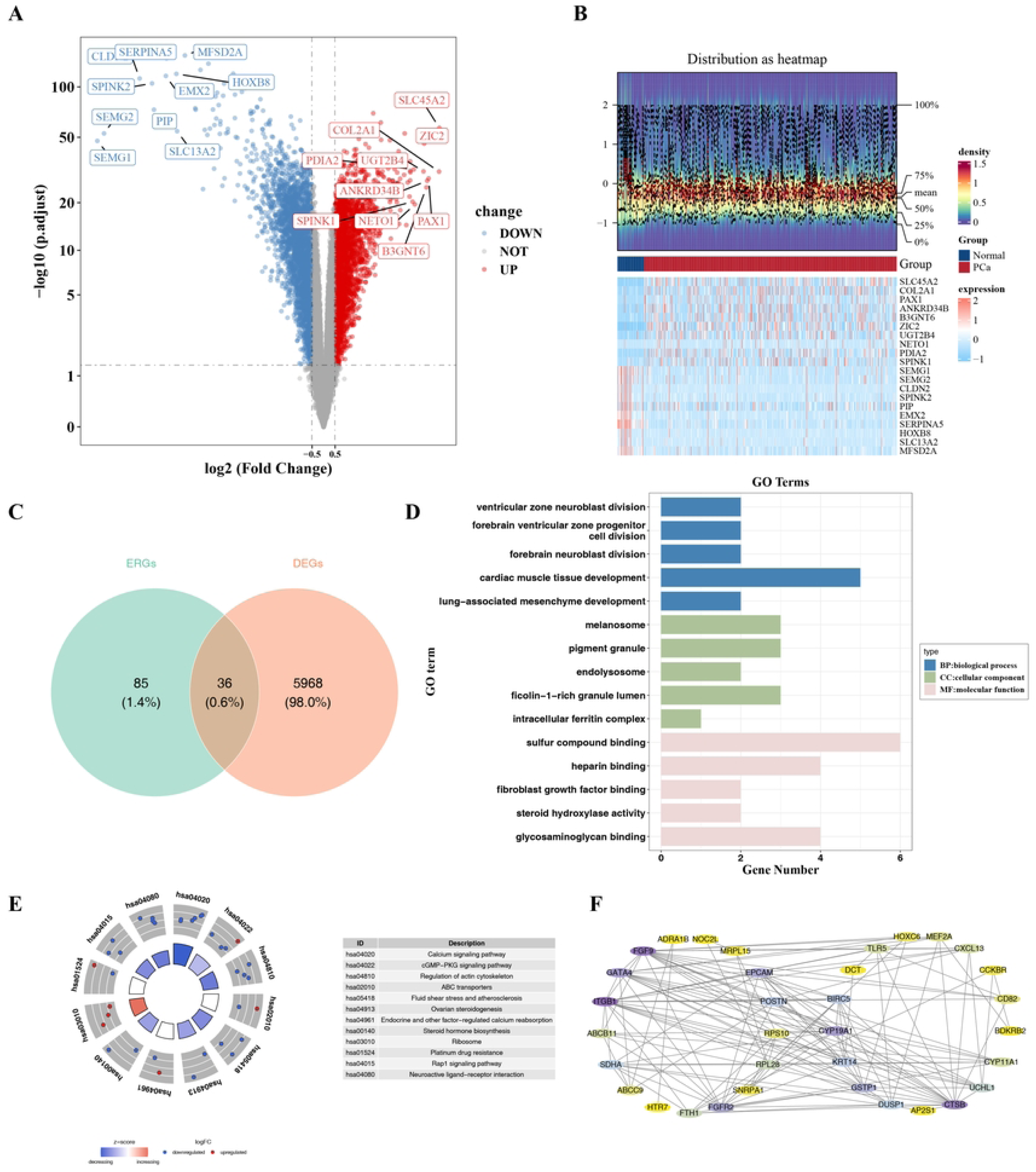
Screening of differentially expressed genes. (A) Volcano plot of DEGs, with red representing upregulation and blue representing downregulation. (B) Heatmap of DEGs, with red indicating higher expression levels and blue indicating lower expression levels. (C) Venn diagram of DEGs and ERGs. (D) GO enrichment analysis plot, where the abscissa represents the gene data enriched in the pathway, and the ordinate represents the corresponding pathway name; the legend on the right indicates the GO enrichment pathway type of the pathway, corresponding to the color of the pathway in the figure. (E) KEGG enrichment analysis plot. The inner left circle is a bar chart, and the height of the bar chart represents the significance of the Term, with higher values indicating greater significance; the outer circle shows a scatter plot of the expression levels of each gene in each Term, with red and blue representing upregulated and downregulated genes respectively; the right side is the KEGG enrichment bar description. (F) PPI network diagram, with colors ranging from light yellow to purple, representing an increase in connectivity from few to many in sequence.

### 3.2 A risk model incorporating four prognostic genes exhibited enhanced predictive accuracy in prostate cancer

A univariate Cox regression showed that 5 candidate prognostic genes exhibited a significant association with OS of PCa (Figure 2A). Furthermore, all 5 genes successfully met the PH assumption test criteria (**S1 Figure**). Following LASSO regression analysis, 4 prognostic genes were identified at optimal lambda 0. 00754352767557399, including NOC2L, RPS10, POSTN, and BIRC5 (Figure 2B). In the training set, 285 PCa samples were divided into HRG (n = 84) and LRG (n = 201) based on the optimal risk score threshold of 6. 004611. The mortality rate of PCa patients progressively increased with higher risk scores (Figure 2C). A substantial survival disparity was observed between HRG and LRG via K-M curves, with the HRG demonstrating a marked reduction in survival probability (P < 0. 0001) (Figure 2D). The AUCs for 1-, 2-, and 3-year were 0. 698, 0. 714, and 0. 764, respectively (Figure 2E). The AUC values were all found to be greater than 0. 6, and the risk model demonstrated a more robust predictive power with PCa prognostic value. In the testing set, 123 PCa patients were categorized into HRG (n = 15) and LRG (n = 108) using the the optimal threshold for the risk score of 6. 408372. In the validation set, a risk score threshold of 20. 57762 was used to classify 248 PCa patients into high-risk (HRG, n = 77) and low-risk groups (LRG, n = 171). Likewise, the results obtained from testing set (Figure 2F-2H) and validation set (Figure 2I-2K) demonstrated the generalizability of the risk model (Figure 2I-2K).

**Figure 2.**
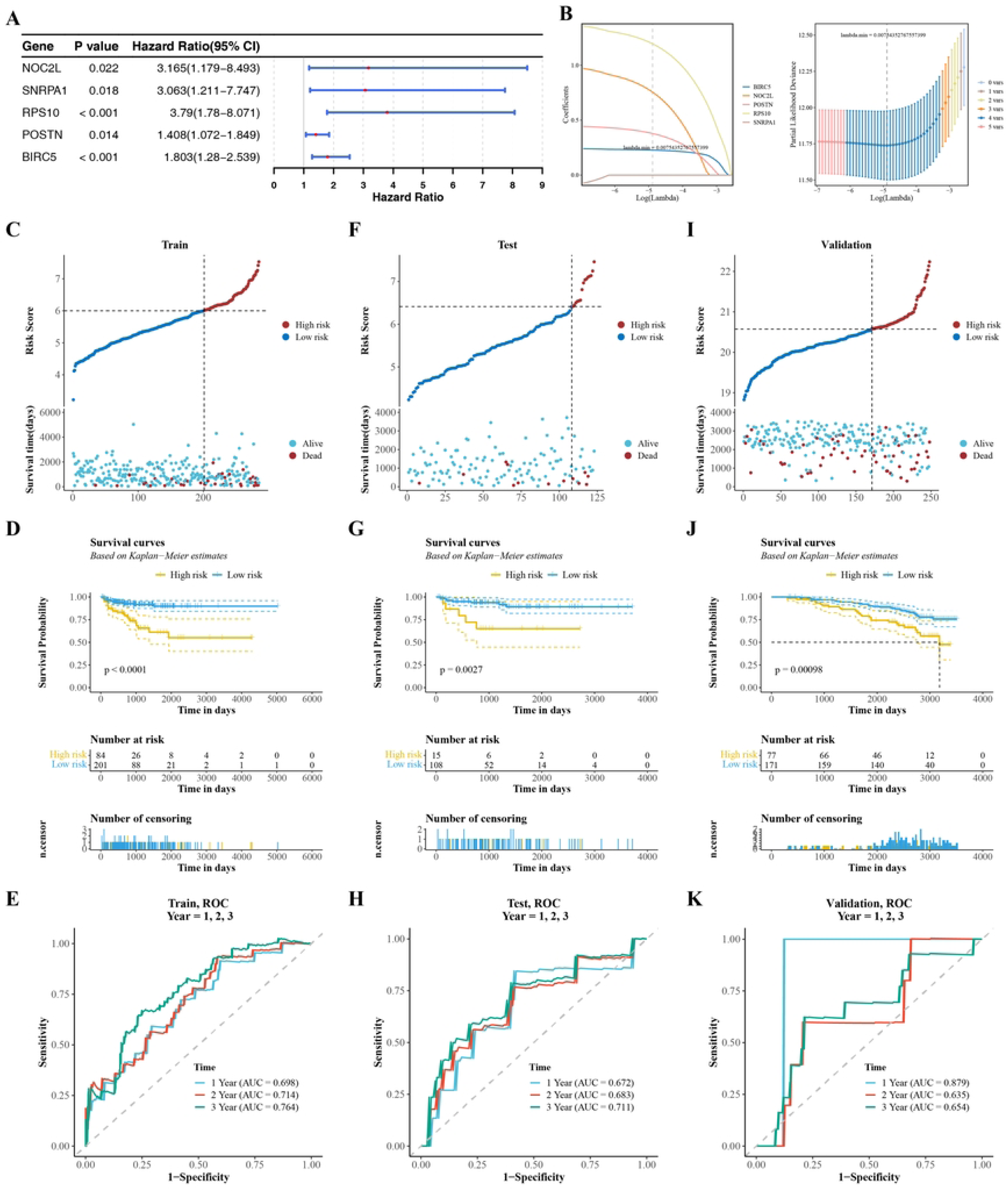
Construction and validation of the risk model. (A) Forest plot of univariate Cox regression analysis. (B) Error plot of Lasso cross - validation and plot of gene coefficients. (C) Analysis plot of risk score and recurrence status in the training set. (D) Construction of Kaplan - Meier recurrence curve in the training set. (E) ROC curves of the model at 1 - year, 2 - year, and 3 - year time points in the training set. (F) Analysis plot of risk score and recurrence status in the validation set. (G) Construction of Kaplan - Meier recurrence curve in the validation set. (H) ROC curves of the model at 1 - year, 2 - year, and 3 - year time points in the validation set. (I) Analysis plot of risk score and recurrence status in the internal validation set. (J) Construction of Kaplan - Meier recurrence curve in the internal validation set. (K) ROC curves of the model at 1 - year, 2 - year, and 3 - year time points in the internal validation set.

### 3.3 A nomogram was developed to predict prostate cancer survival outcomes, utilizing risk score and T stage as independent prognostic indicators

With respect to the clinical characteristics, the data revealed significant variations in risk score among the age, T stage, and N stage groups (Figure 3A). However, no substantial difference was noted in the race.

**Figure 3.**
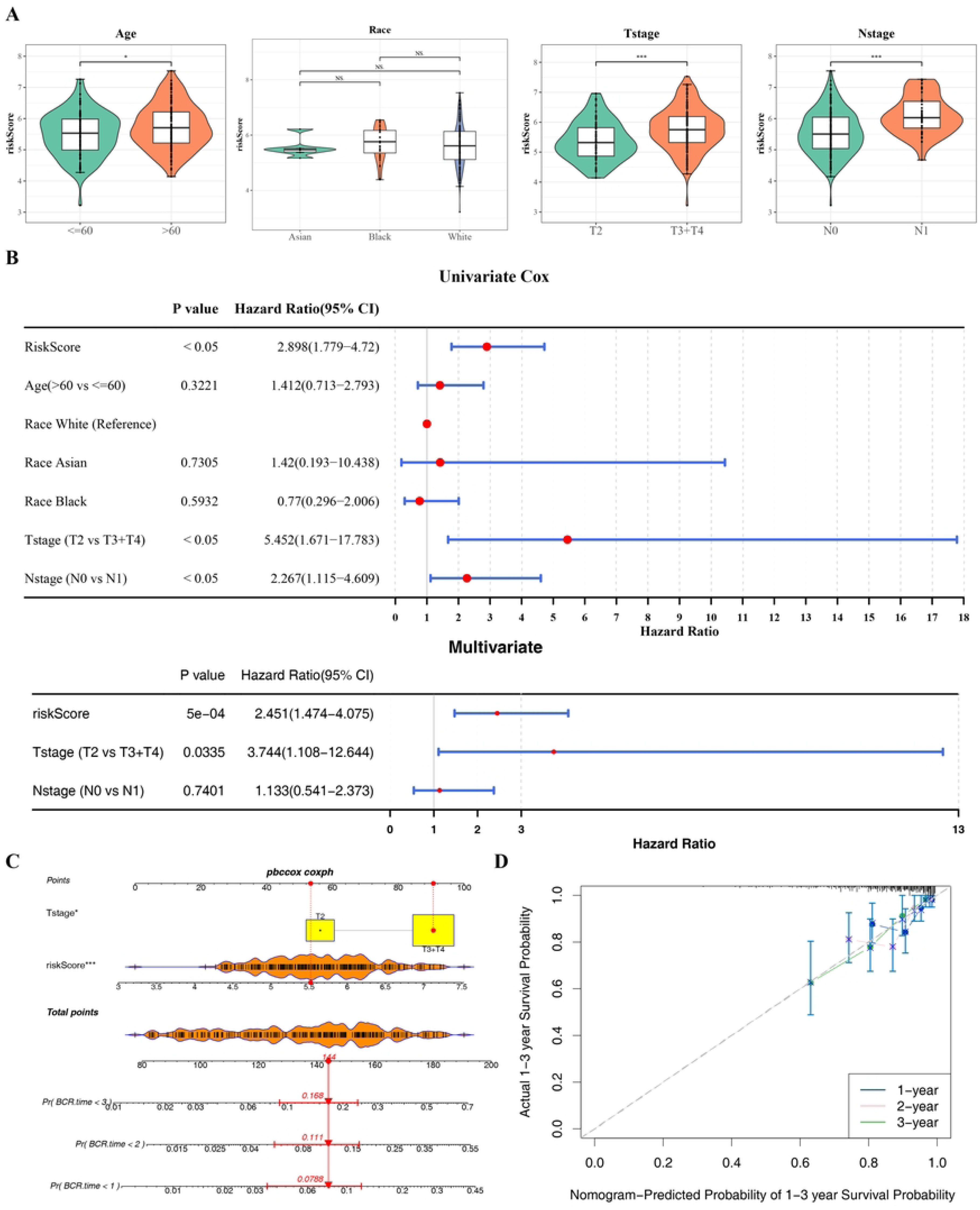
Clinicopathological analysis of prognostic characteristics and construction of Nomogram. (A) Violin plot of differential analysis of risk scores among different clinical feature subtypes. ns represents insignificant, *p<0. 05, **p<0. 01, ***p<0. 001. (B) Independent prognosis - univariate and multivariate Cox analysis. (C) Construction of the Nomogram. Each predictor corresponds to a horizontal line, and different values correspond to different scores.. (D) Calibration curve of the Nomogram.

The univariate Cox regression analysis and proportional hazards assumption test identified risk score, T stage, and N stage as risk factors for overall survival in prostate cancer patients (Figure 3B**, S2 Figure**). In the multivariate Cox analysis for overall survival (OS) in prostate cancer (PCa) patients, both the risk score and T stage were identified as independent prognostic factors (Figure 3B). All 2 independent prognostic factors satisfied the PH assumption test (**S3 Figure**). Then a nomogram incorporating these factors was devised to forecast 1-, 2-, and 3-year OS in PCa (Figure 3C). Furthermore, the calibration curve demonstrated that the actual survival over a period of 1-, 2-, and 3-year exhibited high degree of concordance with the predicted values (Figure 3D).

### 3.4 Exploring the functional and immune infiltration differences between HRG and LRG

GSEA is advantageous method for analyzing biological differences between different risk groups. Then, the signaling pathways between the HRG and LRG were explored by GSEA. GSEA analysis identified 42 signaling pathways with significant differences between the two risk groups, including the ribosome pathway (Figure 4A**, S3 Table**). For HRG and LRG samples, the proportions of 28 kinds of immune cell score were displayed in Figure 4B. A comparative analysis of immune cell infiltration score revealed significant disparities among 7 distinct immune cell types between 2 groups (Figure 4C). Most of the cells exhibiting significant differences demonstrated strong positive correlations with one another (Figure 4D). Effector memory CD8 T cells showed a strong positive correlation with MDSC (r > 0. 80, P < 0. 001). RPS10 demonstrated a strong negative correlation with immature dendritic cells (r < −0. 50, P < 0. 001), while POSTN exhibited a significant positive correlation with effector memory CD8 T cells (r > 0. 50, P < 0. 001) (Figure 4E). The complexity and activity of the tumor microenvironment might make the HRG of tumors more aggressive.

**Figure 4.**
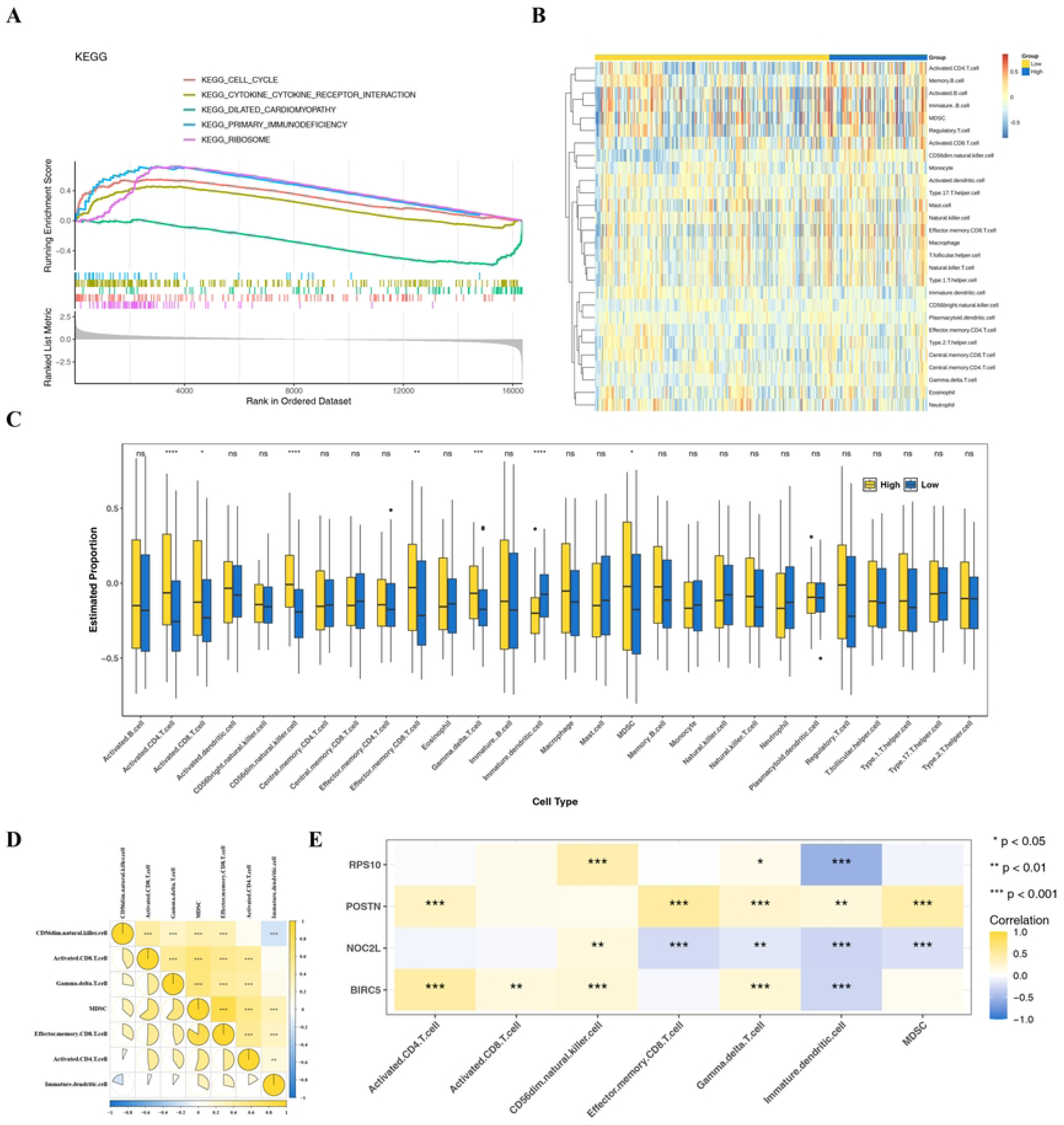
GSEA enrichment and immune infiltration analysis. (A) GSEA-KEGG enrichment analysis of high- and low-risk groups. The curves in the figure represent different pathways. The horizontal axis indicates the rank of genes in the sorted dataset, and the vertical axis represents the running enrichment score. The peak at the top of the curve shows the enrichment level of each gene set in the dataset. The bars below mark the positions of genes, and the density plot at the bottom represents the overall distribution of these genes in the entire dataset. (B) Heatmap of ssGSEA-estimated different cell contents. (C) Expression of immune infiltrating cells in high- and low-risk group samples, ns represents insignificant, *p<0. 05, **p<0. 01, ***p<0. 001, ****p<0. 0001. (D) Heatmap of correlation analysis of differential immune cells. Yellow represents positive correlation, and blue represents negative correlation. ns represents insignificant, *p<0. 05, **p<0. 01, ***p<0. 001. (E) Heatmap of correlation analysis between differential immune cells and prognostic genes. Yellow represents positive correlation, and blue represents negative correlation. ns represents insignificant, *p<0. 05, **p<0. 01, ***p<0. 001.

### 3.5 Significant differences in mutations and chemotherapy drug sensitivity were observed between HRG and LRG

As shown in Figure 5A-5B, the waterfall plots visualized the mutational landscapes of the HRG and LRG. Moreover, missense mutation was identified as the predominant mutation type within the 2 groups. A notable difference in TMB score was identified between the 2 group, with the HRG exhibiting elevated TMB score in comparison to the LRG (Figure 5C).

**Figure 5.**
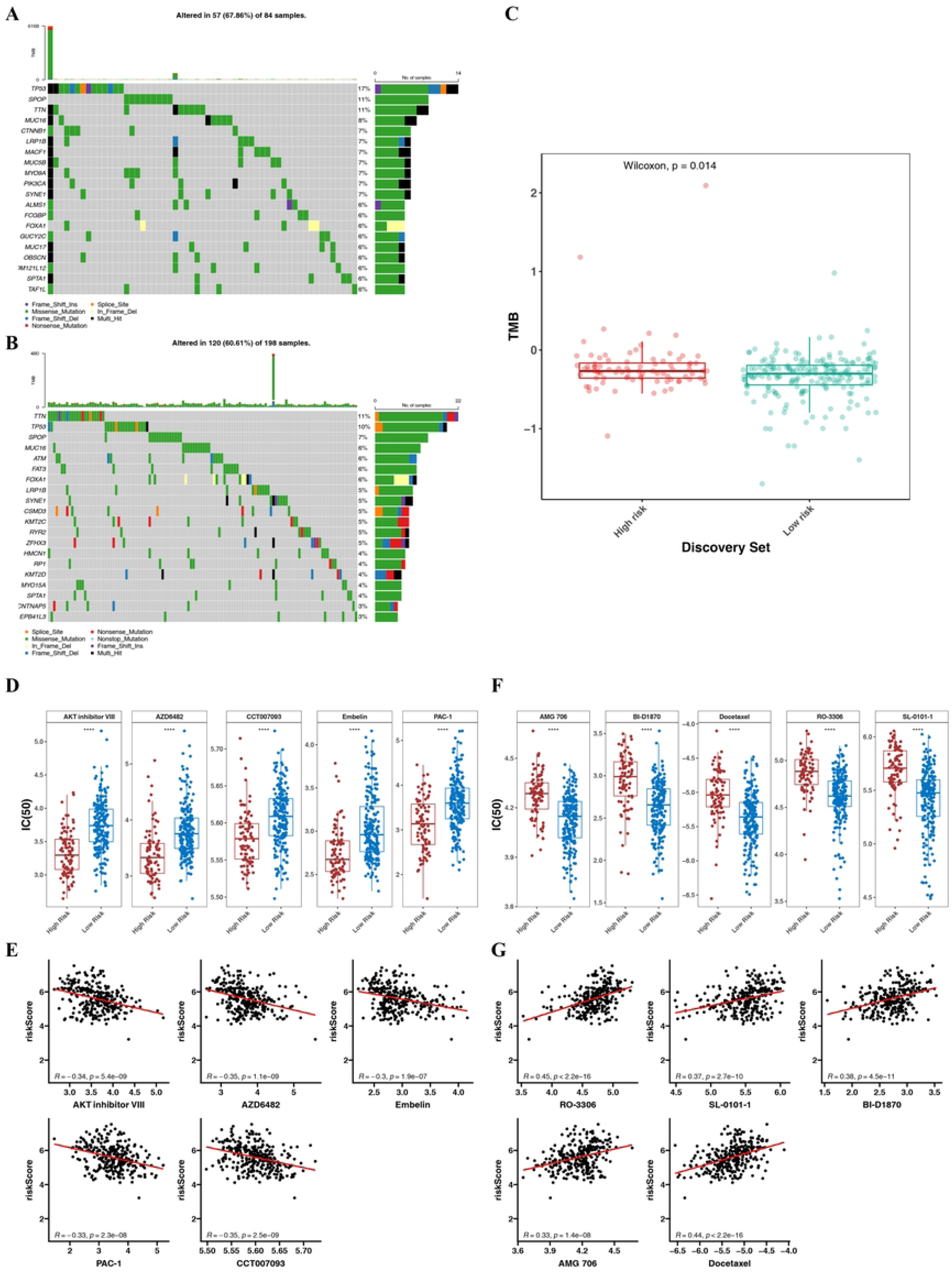
Analysis of somatic mutation signatures and drug sensitivity. Analysis of somatic mutation signatures in the high-risk group (A) and the low-risk group (B). (C) is a diagram of the difference in TMB scores between the high- and low-risk groups. (D) is a diagram of drug sensitivity analysis in the high-risk group, ****p<0. 0001. (E) is a diagram of the correlation analysis between the top 5 sensitive drugs and the risk score in the high-risk group. (F) is a diagram of drug sensitivity analysis in the low-risk group, ****p<0. 0001. (G) is a diagram of the correlation analysis between the top 5 sensitive drugs and the risk score in the low-risk group.

A total of 77 drugs exhibited significant variation between the 2 risk subgroups. Among them, the IC50 values of 27 drugs were significantly lower in the HRG compared to the LRG (**S4 Table**). The top 5 drugs identified were AKT inhibitor VIII, AZD6482, embelin, PAC-1, and CCT007093 (Figure 5D). Moreover, all 5 pharmacological agents demonstrated a substantial inverse relationship with the risk score (Figure 5E). In addition, 70 drugs exhibited significantly higher IC50 values in the HRG compared to the LRG (**S5 Table**). The top 5 drugs identified were RO-3306, SL-0101-1, BI-D1870, AMG 706, and docetaxel (Figure 5F). Moreover, all 5 pharmacological agents exhibited a substantial positive correlation with the risk score (Figure 5G).

### 3.6 Prognostic genes with varying localization were regulated by multiple TFs and miRNAs

Chromosome localization results indicated that NOC2L was located on chromosome 1, RPS10 on chromosome 6, POSTN on chromosome 13, and BIRC5 on chromosome 17 (Figure 6A). In addition, NOC2L and PRS10 proteins were primarily located in the cytosol and nucleus; BIRC5 protein was predominantly found in the cytoskeleton, cytosol, and nucleus; whereas POSTN protein was mainly situated in the extracellular and the Golgi apparatus (Figure 6B). The locational information of prognostic genes facilitated a deeper understanding not only of the distinct impacts on PCa, but also provided a novel perspective for exploring their functional associations.

**Figure 6.**
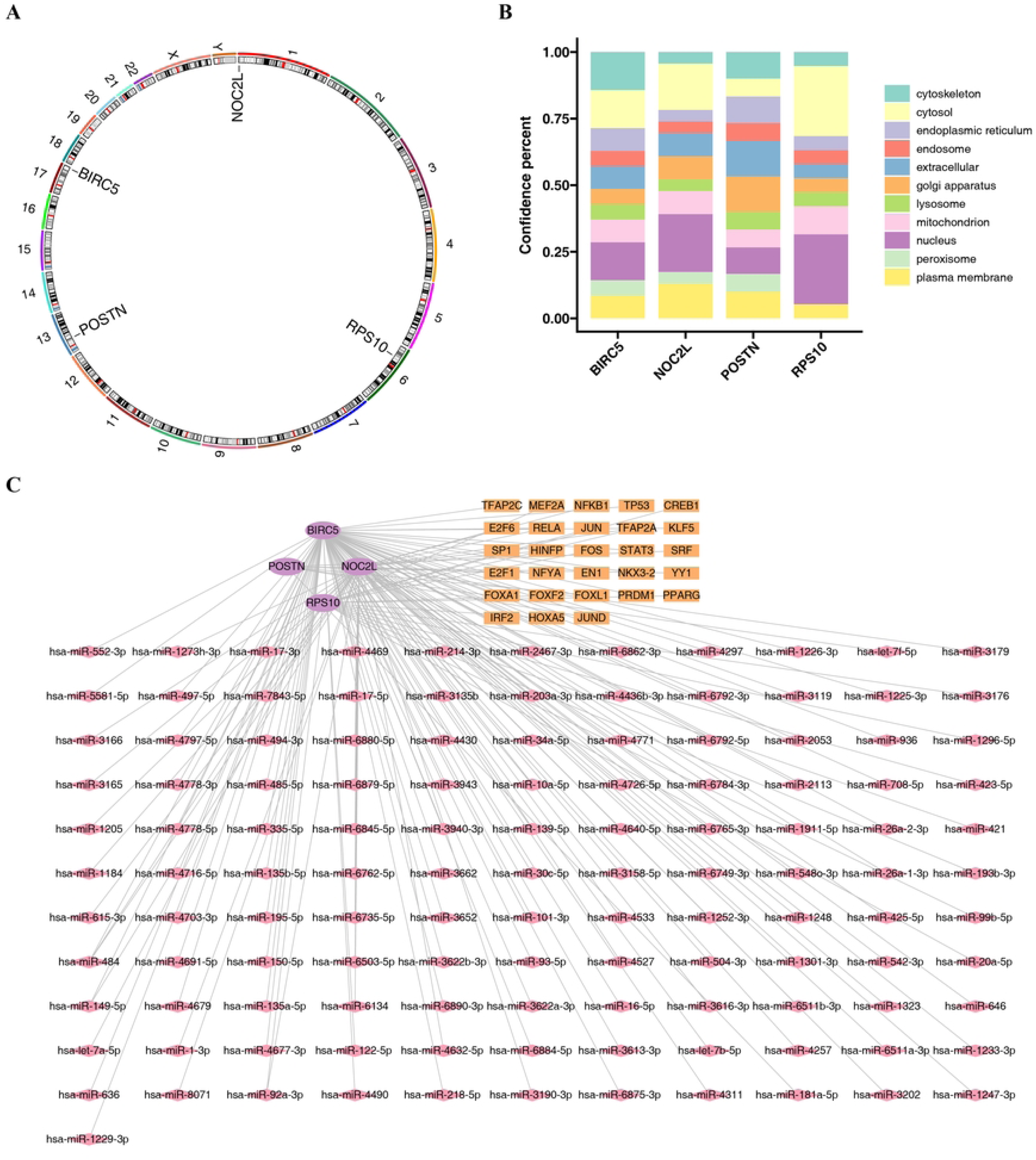
Location analysis and molecular regulatory network analysis. (A) Chromosome location analysis diagram. (B) Subcellular localization analysis of genes. (C) miRNA-mRNA-TF molecular regulatory network. The purple elliptical nodes are prognostic genes, the orange square nodes are TFs, and the pink diamond nodes are miRNAs.

In the TF/miRNA-mRNA network, NOC2L was regulated by 7 TFs and 35 miRNAs, BIRC5 was regulated by 11 TFs and 79 miRNAs, POSTN was regulated by 5 TFs and 2 miRNAs, and RPS10 was regulated by 8 TFs and 10 miRNAs (Figure 6C). In particular, HINFP and RELA were both found to regulate NOC2L and BIRC5, while TFAP2A was identified as a co-regulator of NOC2L and RPS10. Additionally, hsa-miR-484 exerted a regulatory influence on NOC2L, BIRC5, and RPS10.

### 3.7 Epithelial cells, endothelial cells, and T cells were identified as the key cells

First, prior to quality control for the GSE193337 dataset, the initial count comprised 35,224 cells and 24,375 genes. Subsequent to quality control, the cell count was refined to 25,709, while the gene count remained constant at 24,375 (**S4 Figure A**). The top 2,000HVGs were identified, with MTIH, MT1G, and MSMB among the top 10 genes exhibiting the highest variability (**S4 Figure B**). The significance diminished after the 30th principal component, as indicated by the flattening of the scree plot curve at this point. Therefore, we chose the top 30 PCs for additional analysis (**S4 Figure C**). After that, cells were partitioned into 15 cell clusters using the UMAP clustering method (Figure 7A). The 15 cell clusters were categorized into 8 cell types: epithelial cells, smooth muscle cells, fibroblasts, endothelial cells, T cells, B cells, monocytes, and mast cells (Figure 7B-7C). The expression of the 4 prognostic genes was examined in 8 cells, as depicted in Figure 7D. In epithelial cells, endothelial cells, and fibroblasts, the genes NOC2L, POSTIN, and PRS10 demonstrated significant differential expression when comparing the PCa group to the normal group. Similarly, in macrophages and T cells, the genes BIRC5, POSTIN, and PRS10 also showed significant differential expression between 2 groups. Conversely, in B cells, mast cells, and smooth muscle cells, only the PRS10 gene was identified as being significantly differentially expressed between 2 groups. The existing literature indicates that epithelial cells, endothelial cells, and T cells are crucial in the pathogenesis and treatment of PCa. As a result, these 3 cell types were recognized as key cells for further investigation. The outcomes of functional enrichment showed that the key cells were markedly enriched in the material synthesis and metabolic pathways (Figure 7E).

**Figure 7.**
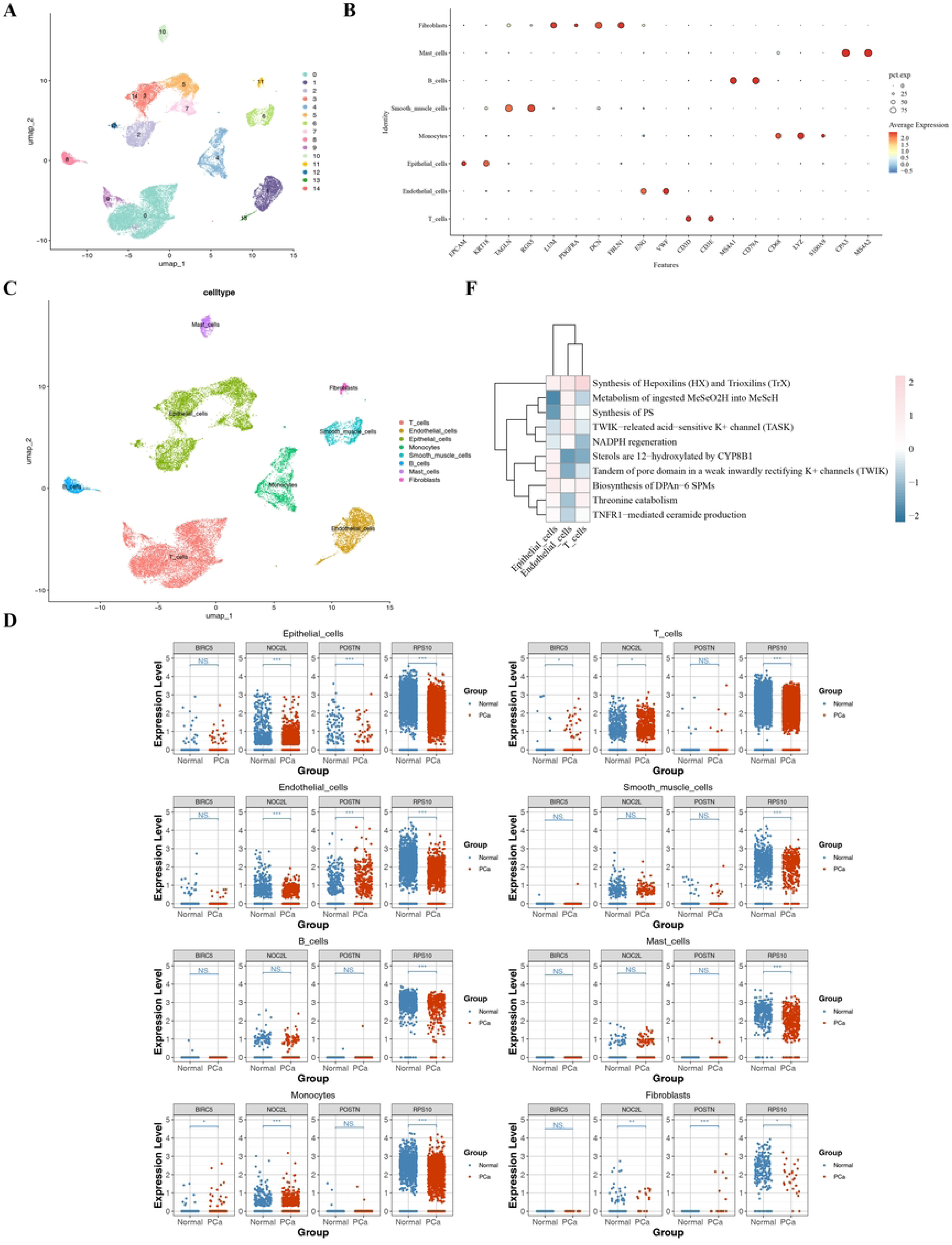
Single - cell analysis. (A) UMAP dimensionality reduction and clustering plot of cells. (B) Bubble plot of the expression of marker genes in different cell populations. (C) Visualization of the annotated cell clusters. (D) Differential analysis of prognostic genes in cells, where NS represents insignificant, *p<0. 05, **p<0. 01, ***p<0. 001, ****p<0. 0001. (E) Differential pathway enrichment analysis.

### 3.8 Exploring the interactions and differentiation of key cells

A comprehensive examination of cellular communication indicated that, in general, the frequency of interactions among the 8 annotated cell types was diminished in the PCa samples relative to the normal samples. Conversely, the intensity of these interactions was found to be heightened in the PCa samples (Figure 8A-8B). Specifically, the interaction of the 3 key cell types with monocytes and smooth muscle cells remained unchanged in both sets of samples. In PCa samples, endothelial cells and fibroblasts exhibited increased interaction, whereas epithelial cells and B cells showed reduced interaction. The interaction between T cells and both epithelial and B cells was enhanced in the PCa samples (Figure 8C-8D). Furthermore, the signals produced by T cells, endothelial cells, and epithelial cells were significantly increased in PCa samples. In contrast, endothelial and epithelial cells exhibited increased signal reception in the PCa samples (Figure 8E-8F). Furthermore, the MIF - (CD74 + CXCR4) signaling axis was identified as a key mediator in the interactions between endothelial cells and B cells in the normal group, while the NAMPT - (ITGA5 + ITGB1) signaling axis was crucial for interactions among endothelial cells in the PCa group. The MIF - (CD74 + CXCR4) signaling axis was identified as a key mediator in the interactions between epithelial cells and B cells in the 2 groups. In contrast, The CCL5 - ACKR1 signaling axis was identified as a key mediator in the interactions between T cells and endothelial cells in both groups (Figure 8G-8H). These findings suggested distinct signaling mechanisms underlying cell communication in different disease contexts.

**Figure 8.**
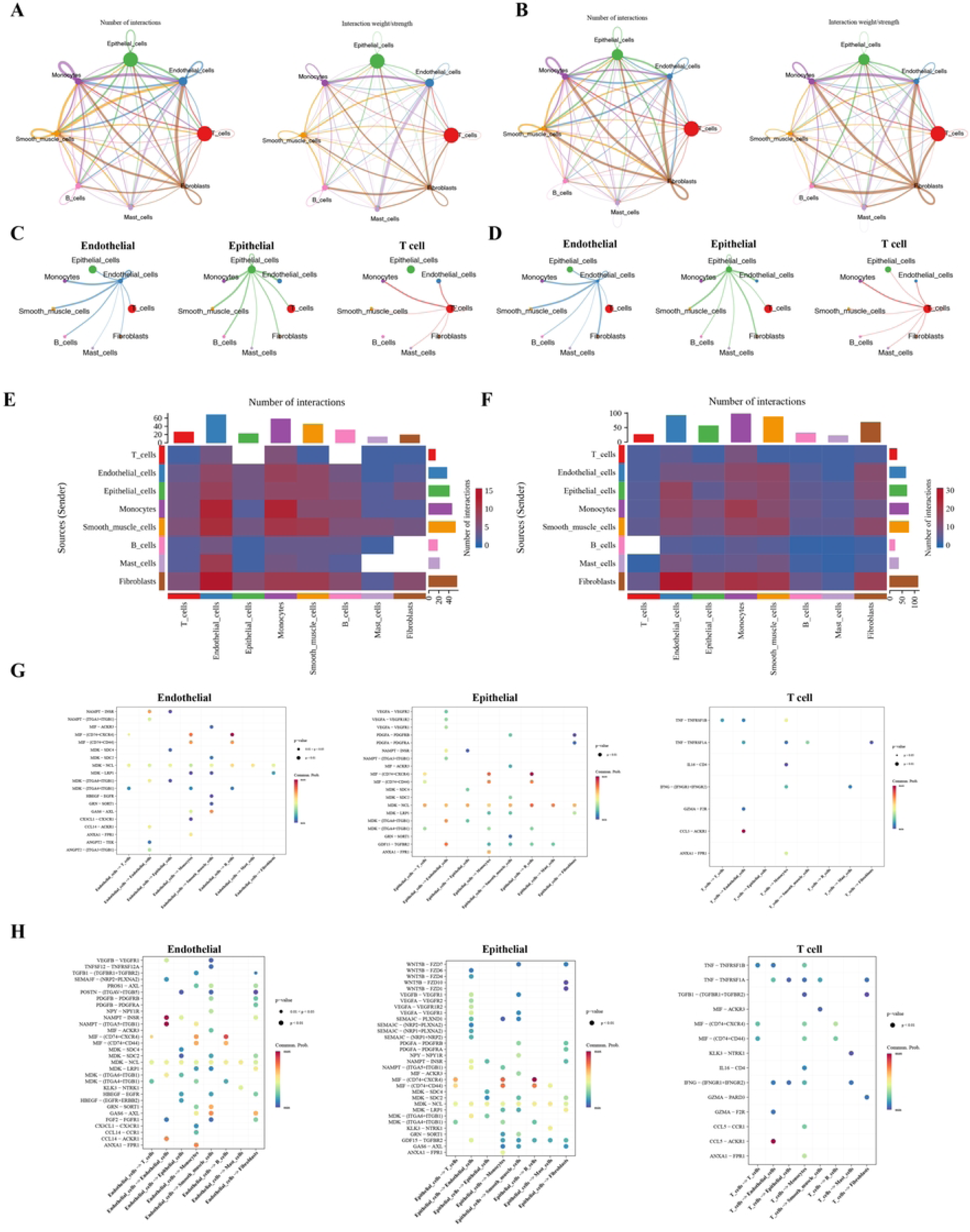
Cell communication analysis. (A-B) Graphs of the number and weight of interactions between cells in the normal and disease groups. The size of the circles of various colors in the periphery represents the number of cells. The larger the circle, the more cells there are. The cells that send out arrows express ligands, and the cells pointed by the arrows express receptors. The more ligand - receptor pairs there are, the thicker the line is. (C-D) Single - network diagrams of key cells Endothelial, Epithelial, and T cells in the normal and disease groups. (G-H) Analysis of the communication probability of ligand - receptor between cells in the normal and disease groups. The abscissa represents the ligand, the arrow points to the receptor, and the ordinate represents the ligand gene and the corresponding receptor gene. The color of the dots represents the communication probability, and the size of the dots represents the significance.

To further assess the cellular differentiation trajectory of the key cells, they were first re-clustered into different cell subgroups. Endothelial cells, epithelial cells, and T cells were divided into 4, 7, and 6 subgroups, respectively (Figure 9A). Pseudo-temporal analysis of endothelial cells, epithelial cells, and T cells differentiation revealed a progressive trajectory from an early (dark red) to a more mature (dark purple) state, and these 3 key cells could be broadly categorized into 5 differentiation states, respectively (Figure 9B). In the course of endothelial cells development, the expression of NOC2L was found to be elevated during the early to middle stages, while BIRC5 demonstrated increased expression specifically during the middle stage. RPS10 showed increased expression in the middle and late stages, while POSTN was more prominently expressed in the early and late stages. In epithelial cell development, NOC2L, BIRC5, RPS10, and POSTN exhibited notable expression levels during the middle and late stages. In T cell development, NOC2L was prominently expressed in the early and middle stages, BIRC5 was mainly expressed in the late stage, while both RPS10 and POSTN exhibited significant expression during the middle and late stages (Figure 9C).

**Figure 9.**
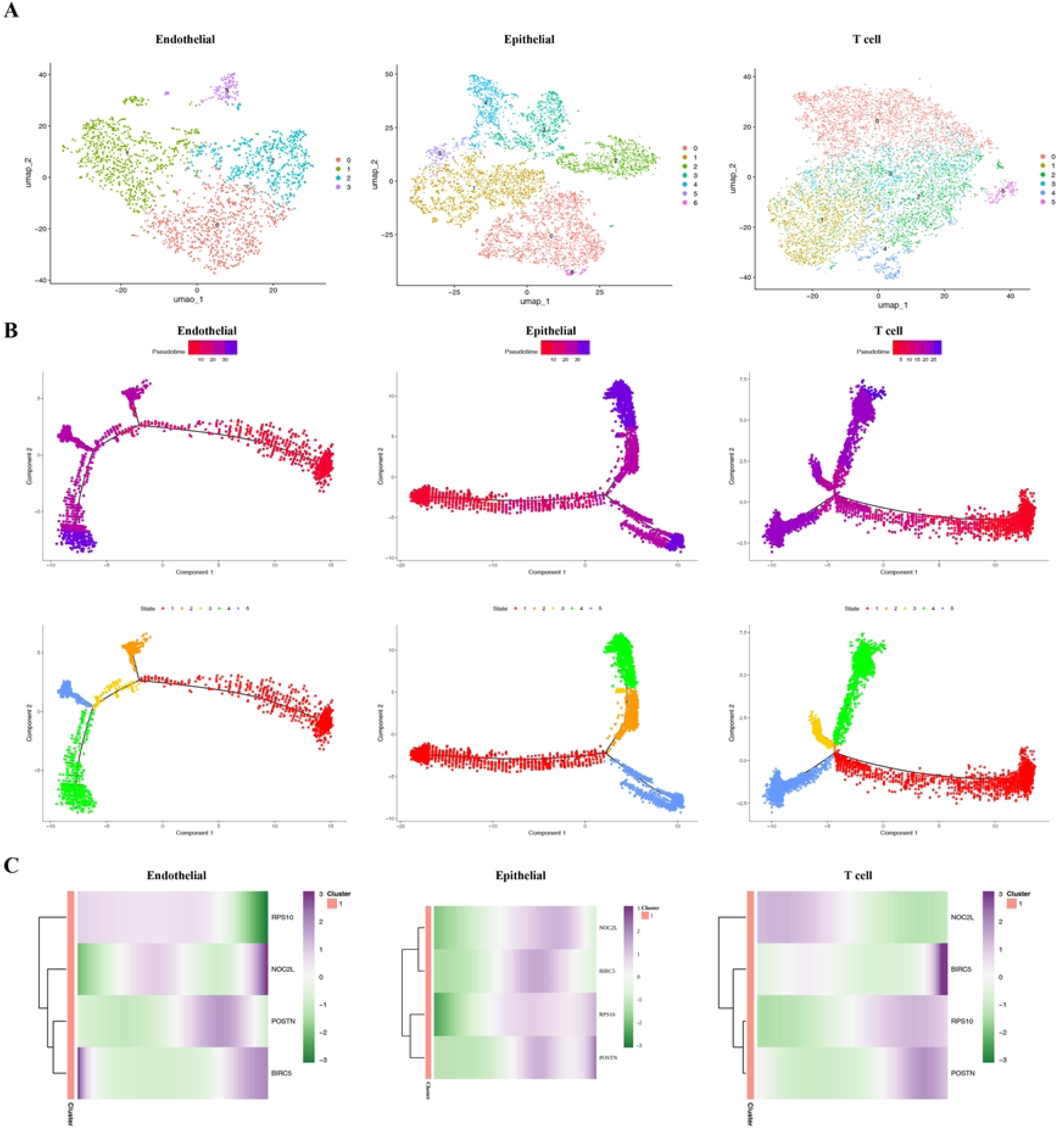
Cell clustering and pseudotemporal sequence analysis. (A) UMAP clustering diagrams of Endothelial, Epithelial, and T cells. (B) Pseudotemporal sequence analysis of Endothelial, Epithelial, and T cells and the expression trends of prognostic genes in cells. (C) Heatmap of the expression of prognostic genes in Endothelial, Epithelial, and T cells at different development times. Different colors in the heatmap represent different expression levels.

## 4. Discussion

Prostate cancer is a common malignant tumor among men worldwide. Its incidence rate is on the rise due to factors such as population aging. It has insidious early symptoms, and in advanced stages, it easily progresses to castration-resistant prostate cancer with treatment resistance, posing a serious threat to men’s health[44]. As a key medium for intercellular communication in the tumor microenvironment, exosomes carry bioactive molecules that can affect the progression of prostate cancer through mechanisms such as regulating immune evasion, promoting metastasis, and inducing drug resistance [45]. The distinct expression profiles of these molecules offer a valuable research avenue for the accurate diagnosis and treatment of prostate cancer, representing a key focus in the ongoing investigation of its pathogenesis and therapeutic approaches. Based on the transcriptomes in GEO and TCGA databases, bioinformatics methods were used to identify exosome-related prognostic genes in PCa, construct prognostic models, and explore their potential molecular regulatory mechanisms. Single-cell analysis was employed to examine prognostic gene expression trends at the cellular level, offering scientific insights and novel targets for PCa prognosis and immunotherapy.

This study utilized machine learning to identify four key prognostic genes—NOC2L, RPS10, POSTN, and BIRC5—by analyzing the intersection of differentially expressed and exosome-related genes. We advanced a precise risk prediction model and clarified the potential roles of these genes in prostate cancer progression.

As a histone acetyltransferase (HAT) inhibitor, NOC2L may be involved in prostate cancer (PCa) progression by disrupting the homeostasis of histone acetylation—histone acetylation, a core process of chromatin remodeling, can affect cell proliferation and differentiation through regulating gene expression [46]. In PCa, the balance between histone deacetylases (HDACs) and HATs is disrupted, leading to abnormal chromatin structure and thereby promoting the proliferation and metastasis of cancer cells [47]; however, no direct evidence of NOC2L in PCa (e. g., tissue expression, functional experiments) has been reported to date, and its specific impact on the HDAC-HAT balance remains to be verified.

The roles of ribosomal proteins (e. g., RPS10, RPL19) in PCa progression extend far beyond protein synthesis: RPS10 can drive the proliferation of PCa cells by enhancing protein synthesis efficiency and is associated with malignant phenotypes [48]; whereas siRNA-mediated RPL19 knockdown can significantly inhibit the invasiveness of PCa cells [48]. Together, these findings suggest that ribosomal proteins may participate in PCa progression by regulating proliferation and invasion.

Periostin (POSTN), an extracellular matrix protein, facilitates prostate cancer invasion and metastasis by promoting ECM remodeling, potentially through SPOCK1-induced epithelial-mesenchymal transition (EMT) and activation of the Wnt/β-catenin pathway. Clinical studies indicate that elevated POSTN expression in prostate cancer (PCa) correlates with unfavorable clinical characteristics and reduced disease-free survival. In metastatic castration-resistant prostate cancer (mCRPC) patients, POSTN expression is linked to increased invasiveness and decreased overall survival. The anti-apoptotic protein BIRC5 (survivin) plays a crucial role in prostate cancer (PCa) progression, therapeutic resistance, and prognosis. High BIRC5 expression is linked to bleomycin resistance, and its inhibition can improve drug efficacy. BIRC5 influences P-glycoprotein expression via the PI3K/Akt/mTOR pathway, contributing to multidrug resistance. Prognostically, elevated BIRC5 levels correlate with poor outcomes in PCa patients, making it a potential prognostic marker. In targeted therapy, siRNA-mediated BIRC5 downregulation enhances selenium’s growth-inhibitory effects on PCa cells, and BIRC5 may be a target of zoledronic acid-induced apoptosis in PCa cells. Notably, the aforementioned molecules may participate in the PCa progression network through pathway crosstalk: for instance, the BIRC5-related PI3K/Akt/mTOR pathway and the POSTN-related Wnt pathway often synergistically promote EMT and drug resistance, while RPS10/RPL19 may provide protein synthesis support for the activation of these pro-carcinogenic pathways; the effect of NOC2L on chromatin remodeling may regulate the transcription of downstream genes in these pathways, but such crosstalk effects still require experimental verification.

GO/KEGG enrichment analysis revealed significant enrichment of the four exosome-related differentially expressed genes in the calcium signaling pathway, steroid hormone synthesis, and actin cytoskeleton regulation. This suggests exosomes may influence PCa progression by modulating intracellular calcium homeostasis and hormone metabolism. For example, abnormal calcium signaling can promote tumor cell migration through activating the NFAT pathway[49–51], while steroid hormone metabolic disorders are closely associated with ADT resistance[52]..

Gene Set Enrichment Analysis (GSEA) identified five significantly enriched pathways—Ribosome, Cell Cycle, Dilated Cardiomyopathy (DCM), Cytokine-Cytokine Receptor Interaction, and Primary Immunodeficiency—when comparing high-risk and low-risk prostate cancer (PCa) groups. The pathways influencing prostate cancer (PCa) progression and prognosis are as follows: Dysregulated ribosome function, marked by increased nucleoli size and number, is linked to poor prognosis. Ribosomal protein L22-like 1 (RPL22L1), part of the 60S ribosomal subunit, promotes PCa cell proliferation, migration, and invasion. Elevated levels of 45S, 28S, 18S, and 5. 8S rRNA in PCa tissues correlate with MYC overexpression, enhancing tumor growth independently of rDNA promoter hypomethylation. Additionally, dysregulated ribosome biogenesis and activation of translation initiation factor eIF4A contribute to the proliferation and drug resistance of castration-resistant prostate cancer (CRPC) by upregulating oncogenes such as AR, AR-V7, and HIF-1α. In the Cell Cycle pathway, dysregulation of regulatory molecules is crucial for prostate cancer (PCa) progression and identifying therapeutic targets. The retinoblastoma protein (Rb) controls the G1-to-S phase transition, while p53 (encoded by TP53) induces cell cycle arrest in response to stress or damage. Its downstream effector, p21 (encoded by CDKN1A), inhibits the cyclin A-CDK1 complex, blocking S-to-G2/M phase progression. High p21 expression is linked to PCa aggressiveness, whereas low expression may promote tumorigenesis. Androgen Deprivation Therapy (ADT) halts PCa cells at the G1 phase by inhibiting cyclin E-CDK2, but cyclin D1 overexpression only partially restores proliferation and does not eliminate androgen dependence. In the DCM pathway, DCM-related genes influence PCa by regulating Lamin A/C, encoded by the LMNA gene, which is overexpressed in PCa. This overexpression activates the PI3K/AKT pathway to enhance proliferation and migration while inhibiting PTEN. Additionally, the lamin A mutant Progerin, present in PCa cells, promotes tumorigenesis by causing DNA damage and genomic instability, sharing pro-aging and pro-carcinogenic mechanisms with DCM-related LMNA mutations. In the Cytokine-Cytokine Receptor Interaction pathway, cytokines in the tumor microenvironment influence prostate cancer (PCa) behavior. CCL4, secreted by macrophages, enhances PCa migration and invasion through androgen receptor (AR) activation. ASC-J9® inhibits AR by reducing the expression of AR-driven oncogenes, such as PSA and TMPRSS2, and blocks CCL4-induced AR activation, thereby curbing PCa proliferation and metastasis. Additionally, IL-6 facilitates PCa proliferation and epithelial-mesenchymal transition via the JAK/STAT3 pathway, with elevated serum IL-6 levels in castration-resistant prostate cancer (CRPC) patients linked to poor prognosis. In the Primary Immunodeficiency pathway, abnormalities in pathway-related genes are associated with PCa initiation and prognosis: the polymorphism at the rs4792800 locus of the TNFSF13B gene correlates with the risk of biochemical recurrence in PCa, and knockdown of TNFSF13B induces cell cycle arrest by regulating the p53 pathway [53]; DNA hypermethylation (corrected from “hypomethylation” in the original text) in PCa induces silencing of CD1 family genes, thereby promoting tumor immune evasion and shaping an immunosuppressive microenvironment [54]. The immunoinfiltration analysis revealed significant differences in immune cell types between high-risk and low-risk prostate cancer groups, specifically identifying variations in activated CD4 and CD8 T cells, CD56 dim natural killer cells, effector memory CD8 T cells, gamma delta T cells, immature dendritic cells, and myeloid-derived suppressor cells (MDSCs). The analysis identified the strongest positive correlation between MDSCs and effector memory CD8 T cells, with POSTN also showing a strong positive correlation with these cells. Conversely, RPS10 demonstrated the strongest negative correlation with immature dendritic cells. The findings indicate that these immune cells could play a role in regulating PCa progression. Enhancing comprehension of their interaction mechanisms may facilitate the creation of innovative immunotherapeutic strategies, improving PCa patient prognosis.

High infiltration of myeloid-derived suppressor cells (MDSCs) promotes immune escape by primarily suppressing T-cell and NK-cell functions [55, 56]. Additionally, MDSCs protect proliferating tumor cells against senescence [57]. MDSCs contribute to castration-resistant prostate cancer (CRPC) progression through interleukin-23 (IL-23) secretion [58]. Multiple studies confirm that targeting MDSCs or neutralizing their recruitment cytokines effectively inhibits tumor progression in various PCa models [58–61]. Furthermore, combining immune checkpoint blockade (ICB) therapy with MDSC-targeted treatments demonstrates significant synergistic therapeutic effects in both primary and metastatic CRPC [62, 63].

Analysis of drug sensitivity showed notable efficacy differences in 77 chemotherapeutic agents between high-risk and low-risk prostate cancer subgroups. Patients in the low-risk group demonstrated increased sensitivity to the inhibitors RO. 3306 (CDK1), SL. 0101. 1 (AKT), BI-D1870 (RSK), AMG-706 (VEGFR), and the chemotherapy drug Docetaxel, with a positive correlation observed between risk scores and drug sensitivity. In contrast, high-risk group patients exhibited higher sensitivity to AKT inhibitor VIII, AZD6482 (PI3K inhibitor), Embelin (XIAP inhibitor), PAC-1 (procaspase-3 activator), and CCT007093 (CHK1 inhibitor), where sensitivity was negatively correlated with risk scores. This divergence provides a theoretical basis for risk-stratified treatment strategies.

Docetaxel, a microtubule stabilizer, binds to β-tubulin subunits to prevent microtubule depolymerization, induces G2/M-phase cell cycle arrest, and reduces the expression of anti-apoptotic proteins Bcl-2/Bcl-xL, thereby facilitating tumor cell apoptosis [64, 65].. Docetaxel is a primary treatment for metastatic castration-resistant prostate cancer (CRPC). Phase III clinical trials (TAX327) confirmed its extension of overall survival (median survival increased by 2. 4 months), with mechanisms and efficacy validated across multiple studies [66–68]. Notably, other low-risk-sensitive drugs like AMG-706 inhibit tumor angiogenesis to block nutrient supply, while high-risk-sensitive agents like AZD6482 suppress proliferation by blocking the PI3K/AKT pathway. These mechanisms have been reported in PCa (e. g., AZD6482 efficacy in PTEN-deficient CRPC)[59].

This study employed single-cell RNA sequencing (scRNA-seq) for an integrated analysis to clarify the cellular and molecular mechanisms driving prostate cancer progression. The results revealed that PCa progression is primarily regulated by three core cell types: epithelial cells, endothelial cells, and T cells. Meanwhile, the study clarified the spatiotemporal expression patterns of exosome-related prognostic genes, uncovered exosome-mediated intercellular communication, and identified the immune characteristics of high-risk PCa, thereby providing key evidence for understanding the pathological mechanisms of PCa and its clinical translation.

Specifically, epithelial cells drive intratumoral and intertumoral heterogeneity through interactions with stromal cells, which is a core feature of PCa pathogenesis [69, 70]. These epithelial cells display varied high androgen receptor (AR) signal expression, linked to castration-resistant prostate cancer (CRPC) development, aligning with findings that low-risk prostate cancer is sensitive to AR pathway inhibitors [38]. The exosome-related prognostic gene POSTN is notably upregulated during the middle and late stages of epithelial cell differentiation, exhibiting spatiotemporal colocalization with the epithelial-mesenchymal transition (EMT) process. This further verifies the core role of POSTN in the PCa invasion-metastasis cascade, which is consistent with previous reports that POSTN promotes tumor invasion by remodeling the extracellular matrix [38].

Tumor endothelial cells exhibit high heterogeneity, which directly affects PCa progression and can serve as a potential therapeutic target [71, 72]. The RAECs subpopulation, identified as ‘tip cells and immature endothelial cells,’ is crucial in tumor angiogenesis and its abundance correlates with poor survival in PCa patients [73]. Another exosome-related prognostic gene, BIRC5, is highly expressed in the late stage of vascular endothelial cell differentiation and is highly consistent with the pseudotemporal trajectory of angiogenesis, suggesting that BIRC5 can enhance the metastatic capacity of PCa by promoting tumor vascular maturation. Furthermore, endothelial cells express immunosuppressive molecules such as PD-L1 and are involved in tumor immune evasion, which explains why high-risk PCa patients are sensitive to immunomodulators like Embelin (consistent with the previous drug sensitivity analysis results).

T cells, central to the anti-tumor immune response, undergo a life cycle from activation to effector function, governed by genetic programs [73]. Intratumoral T cell infiltration is associated with the clinical outcome of PCa; in particular, CD8⁺T cells can directly exert anti-tumor effects by killing malignant cells [74, 75]. In the prostate cancer microenvironment, T cell exhaustion frequently occurs due to myeloid-derived suppressor cell infiltration or immunosuppressive factors like IL-10 [76–79]. This exhaustion impairs the anti-tumor immune effect, suggesting that combined immune checkpoint blockade (ICB) and microenvironment regulation (e. g., targeting the CCL20-CCR6 axis) are required to restore T cell function [80–83]. This mechanism is also consistent with the phenomenon that high-risk PCa is sensitive to PAC-1, an activator of the apoptotic pathway.

Further analysis of the intercellular communication network showed that ligand-receptor interactions among core cells mainly rely on exosome mediation (in line with the core focus of this study on “exosome-related genes”). Specifically, endothelial cells secrete nicotinamide phosphoribosyltransferase (NAMPT) via exosomes, which activates the energy metabolism pathway in epithelial cells [84], thereby enhancing the proliferation and metastatic potential of epithelial cells and establishing a direct link between endothelial cell function and epithelial cell-driven PCa progression. In turn, epithelial cells secrete macrophage migration inhibitory factor (MIF) via exosomes. MIF binding to CD74/CXCR4 receptors on T cells inhibits their anti-tumor activity and recruits MDSCs, creating an immunosuppressive microenvironment that facilitates PCa immune evasion [85].

Notably, high-risk PCa exhibits a significant feature of “immune activation-suppression imbalance”: activated CD4⁺/CD8⁺T cells (which promote anti-tumor effects) and MDSCs (which promote immunosuppression) infiltrate simultaneously. This imbalance serves as a key mechanism for tumor immune evasion—activated T cells cannot effectively exert anti-tumor effects due to the MDSC-mediated inhibitory effect. At the same time, high-risk PCa also shows increased tumor mutation burden (TMB) and elevated TP53 mutation load. These characteristics collectively suggest the potential value of combined immunotherapy (e. g., ICB combined with MDSC-targeted therapy) for high-risk patients.

In conclusion, through single-cell analysis, this study identified three core mechanisms underlying PCa progression: (1) POSTN in epithelial cells and BIRC5 in endothelial cells regulate PCa invasion-metastasis and angiogenesis, respectively, through stage-specific expression; (2) the exosome-mediated NAMPT and MIF pathways coordinate intercellular functions to drive disease progression; and (3) immune imbalance in high-risk PCa is the key to tumor evasion. These findings offer direct evidence of exosome-mediated intercellular communication in PCa and establish a theoretical basis for precision therapies targeting the MIF-CD74 axis or NAMPT pathway.

This study identified four exosome-related prognostic genes in prostate cancer (PCa), namely NOC2L, RPS10, POSTN, and BIRC5. The findings indicate a strong link between these genes and the onset and development of PCa, especially in the context of immune cell infiltration regulation. A risk model developed based on these four prognostic genes demonstrated robust and stable predictive capability for PCa prognosis. Analysis of the distribution and expression patterns of these genes in single-cell datasets further identified endothelial cells, epithelial cells, and T cells as key cellular players in PCa pathogenesis. These findings provide a novel theoretical foundation and direction for future exploration of the specific mechanisms through which these genes drive PCa progression.

Nonetheless, this research possesses certain limitations. First, the precise molecular mechanisms underlying these prognostic genes in PCa remain unexplored. Additionally, while the constructed risk model exhibits strong prognostic predictive power, its practical utility in clinical applications and therapeutic efficacy requires robust evidence-based medical validation. Future research should aim to clarify the functional roles of these genes and explore their therapeutic applications to validate their clinical significance.

## Glossary

| Full Name | Abbreviation |
| --- | --- |
| Prostate cancer | PCa |
| androgen deprivation therapy | ADT |
| castration-resistant prostate cancer | CRPC |
| cancer-associated fibroblasts | CAFs |
| benign prostatic hyperplasia | BPH |
| Prostate-specific antigen | PSA |
| tumor microenvironment | TME |
| Gene Expression Omnibus | GEO |
| The Cancer Genome Atlas | TCGA |
| Differentially Expressed Genes | DEGs |
| estrogen-related genes | ERGs |
| protein-protein interaction | PPI |
| overall survival | OS |
| hazard ratio | HR |
| Least Absolute Shrinkage and Selection Operator | LASSO |
| high-risk | HRG |
| low-risk | LRG |
| gene set enrichment analysis | GSEA |
| gene ontology | GO |
| Kyoto Encyclopedia of Genes and Genomes | KEGG |
| tumor mutation burden | TMB |
| transcription factors | TFs |
| microRNAs | miRNAs |
| highly variable genes | HVGs |
| histone deacetylases | HDACs |
| Periostin | POSTN |
| epithelial-mesenchymal transition | EMT |
| metastatic castration-resistant prostate cancer | mCRPC |
| retinoblastoma protein | Rb |
| myeloid-derived suppressor cells | MDSCs |
| immune checkpoint blockade | ICB |
| single-cell RNA sequencing | scRNA-seq |
| nicotinamide phosphoribosyltransferase | NAMPT |
| migration inhibitory factor | MIF |

## Data Availability

The data underlying the results presented in the study are available from public omics databases and published literature, with detailed access information and relevant specifications provided as follows:RNA-sequencing (RNA-seq) data, somatic mutation data, clinical characteristics, and survival information of prostate cancer (PCa) patients were downloaded from The Cancer Genome Atlas (TCGA) database (URL: https://portal.gdc.cancer.gov/) on March 20, 2025. The TCGA-PCa dataset includes 51 normal tissue samples and 484 PCa tissue samples, among which 408 PCa samples with available survival information were randomly divided into a training set (285 samples) and a testing set (123 samples) at a 7:3 ratio.For external validation, 248 PCa tissue samples with complete survival details and gene expression data were selected from the GSE116918 dataset in the Gene Expression Omnibus (GEO) database (URL: https://www.ncbi.nlm.nih.gov/geo/) based on the GPL25318 platform. Single-cell RNA sequencing (scRNA-seq) data were retrieved from the GEO database (URL: https://www.ncbi.nlm.nih.gov/geo/) with the accession number GSE193337, which was generated using the GPL20301 sequencing platform and contains 4 PCa samples and 4 normal samples. Additionally, 121 exosome-related genes (ERGs) were identified from published literature (PMID: 34659224), and the detailed list of these genes is provided in Supporting Information Table S1.All public omics data were accessed and analyzed in strict compliance with the data usage policies of the TCGA and GEO databases.

## Acknowledgments

The author would like to express sincere gratitude to all individuals, institutions, and parties involved in supporting this research.

## Ethics approval

Not applicable.

## Consent to participate

Not applicable.

## Consent for publication

Not applicable.

## Availability of data and materials

The datasets analysed during the current study are available in the TCGA database, [https://portal.gdc.cancer.gov/], including TCGA-PCa dataset; and the GEO database, [https://www.ncbi.nlm.nih.gov/geo/],. including GSE116918 and GSE193337 dataset.

## Author contributions

The author was responsible for study conception and design, data collection, data analysis and chart organization, manuscript drafting, review and revision of important intellectual content, and final approval of the published version.

## Supporting information

**S1 Figure** Residual plots of PH assumption tests for prognosis - related genes (BIRC5, NOC2L, POSTN, RPS10, SNRPA1).

**S2 Figure** Univariate PH assumption test for independent prognosis.

**S3 Figure** Multivariate PH assumption test for independent prognosis.

**S4 Figure** Single - cell data processing. (A) Distribution plots before and after single - cell data quality control. (B) Screening of highly variable genes, with highly variable genes shown in red. (C) PCA dimensionality reduction plot and scree plot.

**S1 Table** Information of ERGs.

**S2 Table** Results of GO enrichment analysis.

**S3 Table** Results of GSEA enrichment analysis in high- and low-risk groups.

**S4 Table** Half maximal inhibitory concentration values of drugs in the HRG group.

**S5 Table** Half maximal inhibitory concentration values of drugs in the HRG group.

## References

1. Wasim S, Lee SY, Kim J. Complexities of Prostate Cancer. Int J Mol Sci. 2022;23.

2. Xia C, Dong X, Li H, Cao M, Sun D, He S, et al. Cancer statistics in China and United States, 2022: profiles, trends, and determinants. Chin Med J (Engl). 2022;135: 584–590.

3. Siegel RL, Miller KD, Wagle NS, Jemal A. Cancer statistics, 2023. CA Cancer J Clin. 2023;73: 17–48.

4. Sung H, Ferlay J, Siegel RL, Laversanne M, Soerjomataram I, Jemal A, et al. Global Cancer Statistics 2020: GLOBOCAN Estimates of Incidence and Mortality Worldwide for 36 Cancers in 185 Countries. CA Cancer J Clin. 2021;71: 209–249.

5. Sandhu S, Moore CM, Chiong E, Beltran H, Bristow RG, Williams SG. Prostate cancer. Lancet. 2021;398: 1075–1090.

6. Li J, Wang X, Xue L, He Q. Exploring the therapeutic mechanism of curcumin in prostate cancer using network pharmacology and molecular docking. Heliyon. 2024;10: e33103.

7. Ortega A, Martinez-Arroyo O, Forner MJ, Cortes R. Exosomes as Drug Delivery Systems: Endogenous Nanovehicles for Treatment of Systemic Lupus Erythematosus. Pharmaceutics. 2020;13: 3.

8. Tenchov R, Sasso JM, Wang X, Liaw WS, Chen CA, Zhou QA. Exosomes─Nature’s Lipid Nanoparticles, a Rising Star in Drug Delivery and Diagnostics. ACS Nano. 2022;16: 17802–17846.

9. Akoto T, Saini S. Role of Exosomes in Prostate Cancer Metastasis. Int J Mol Sci. 2021;22: 3528.

10. Wu Y, Wang X, Zeng Y, Liu X. Exosomes are the mediators between the tumor microenvironment and prostate cancer (Review). Exp Ther Med. 2024;28: 439.

11. Liu Z, Lin Z, Jiang M, Zhu G, Xiong T, Cao F, et al. Cancer-associated fibroblast exosomes promote prostate cancer metastasis through miR-500a-3p/FBXW7/HSF1 axis under hypoxic microenvironment. Cancer Gene Ther. 2024;31: 698–709.

12. Zhang H, Li M, Zhang J, Shen Y, Gui Q. Exosomal Circ-XIAP Promotes Docetaxel Resistance in Prostate Cancer by Regulating miR-1182/TPD52 Axis. Drug Des Devel Ther. 2021;15: 1835–1849.

13. Gaglani S, Gonzalez-Kozlova E, Lundon DJ, Tewari AK, Dogra N, Kyprianou N. Exosomes as A Next-Generation Diagnostic and Therapeutic Tool in Prostate Cancer. Int J Mol Sci. 2021;22: 10131.

14. Hamid Y, Rabbani RD, Afsara R, Nowrin S, Ghose A, Papadopoulos V, et al. Exosomal Liquid Biopsy in Prostate Cancer: A Systematic Review of Biomarkers for Diagnosis, Prognosis, and Treatment Response. Int J Mol Sci. 2025;26: 802.

15. Logozzi M, Angelini DF, Giuliani A, Mizzoni D, Di Raimo R, Maggi M, et al. Increased Plasmatic Levels of PSA-Expressing Exosomes Distinguish Prostate Cancer Patients from Benign Prostatic Hyperplasia: A Prospective Study. Cancers (Basel). 2019;11: 1449.

16. Bhagirath D, Yang TL, Bucay N, Sekhon K, Majid S, Shahryari V, et al. microRNA-1246 Is an Exosomal Biomarker for Aggressive Prostate Cancer. Cancer Res. 2018;78: 1833–1844.

17. Zhao J, Shi Y, Cao G. The Application of Single-Cell RNA Sequencing in the Inflammatory Tumor Microenvironment. Biomolecules. 2023;13: 344.

18. Ding S, Chen X, Shen K. Single-cell RNA sequencing in breast cancer: Understanding tumor heterogeneity and paving roads to individualized therapy. Cancer Commun (Lond). 2020;40: 329–344.

19. Jovic D, Liang X, Zeng H, Lin L, Xu F, Luo Y. Single-cell RNA sequencing technologies and applications: A brief overview. Clin Transl Med. 2022;12: e694.

20. Qiu P, Guo Q, Yao Q, Chen J, Lin J. Characterization of Exosome-Related Gene Risk Model to Evaluate the Tumor Immune Microenvironment and Predict Prognosis in Triple-Negative Breast Cancer. Front Immunol. 2021;12: 736030.

21. Love MI, Huber W, Anders S. Moderated estimation of fold change and dispersion for RNA-seq data with DESeq2. Genome Biol. 2014;15: 550.

22. Gustavsson EK, Zhang D, Reynolds RH, Garcia-Ruiz S, Ryten M. ggtranscript: an R package for the visualization and interpretation of transcript isoforms using ggplot2. Bioinformatics. 2022;38: 3844–3846.

23. Hu K. Become Competent in Generating RNA-Seq Heat Maps in One Day for Novices Without Prior R Experience. Methods Mol Biol. 2021;2239: 269–303.

24. Wu T, Hu E, Xu S, Chen M, Guo P, Dai Z, et al. clusterProfiler 4. 0: A universal enrichment tool for interpreting omics data. Innovation (Camb). 2021;2: 100141.

25. Shannon P, Markiel A, Ozier O, Baliga NS, Wang JT, Ramage D, et al. Cytoscape: a software environment for integrated models of biomolecular interaction networks. Genome Res. 2003;13: 2498–2504.

26. Lei J, Qu T, Cha L, Tian L, Qiu F, Guo W, et al. Clinicopathological characteristics of pheochromocytoma/paraganglioma and screening of prognostic markers. J Surg Oncol. 2023;128: 510–518.

27. Tamiru B, Soromessa T, Warkineh B, Legesse G, Belina M. Woody species composition and community types of Hangadi Watershed, Guji Zone, Ethiopia. BMC Ecol Evol. 2021;21: 225.

28. Friedman J, Hastie T, Tibshirani R. Regularization Paths for Generalized Linear Models via Coordinate Descent. J Stat Softw. 2010;33: 1–22.

29. Navarro G, Gómez-Autet M, Morales P, Rebassa JB, Llinas Del Torrent C, Jagerovic N, et al. Homodimerization of CB(2) cannabinoid receptor triggered by a bivalent ligand enhances cellular signaling. Pharmacol Res. 2024;208: 107363.

30. Hänzelmann S, Castelo R, Guinney J. GSVA: gene set variation analysis for microarray and RNA-seq data. BMC Bioinformatics. 2013;14: 7.

31. Liu ZY, Huang RH. Integrating single-cell RNA-sequencing and bulk RNA-sequencing data to explore the role of mitophagy-related genes in prostate cancer. Heliyon. 2024;10: e30766.

32. Robles-Jimenez LE, Aranda-Aguirre E, Castelan-Ortega OA, Shettino-Bermudez BS, Ortiz-Salinas R, Miranda M, et al. Worldwide Traceability of Antibiotic Residues from Livestock in Wastewater and Soil: A Systematic Review. Animals (Basel). 2021;12:60.

33. Mayakonda A, Lin DC, Assenov Y, Plass C, Koeffler HP. Maftools: efficient and comprehensive analysis of somatic variants in cancer. Genome Res. 2018;28: 1747–1756.

34. Geeleher P, Cox N, Huang RS. pRRophetic: an R package for prediction of clinical chemotherapeutic response from tumor gene expression levels. PLoS One. 2014;9: e107468.

35. Zhang H, Meltzer P, Davis S. RCircos: an R package for Circos 2D track plots. BMC Bioinformatics. 2013;14: 244.

36. Satija R, Farrell JA, Gennert D, Schier AF, Regev A. Spatial reconstruction of single-cell gene expression data. Nat Biotechnol. 2015;33: 495–502.

37. Liu W, Wang M, Wang M, Liu M. Single-cell and bulk RNA sequencing reveal cancer-associated fibroblast heterogeneity and a prognostic signature in prostate cancer. Medicine (Baltimore). 2023;102: e34611.

38. Song H, Weinstein HNW, Allegakoen P, Wadsworth MH, 2nd, Xie J, Yang H, et al. Single-cell analysis of human primary prostate cancer reveals the heterogeneity of tumor-associated epithelial cell states. Nat Commun. 2022;13: 141.

39. Ji S, Wu W, Jiang Q. Crosstalk between Endothelial Cells and Tumor Cells: A New Era in Prostate Cancer Progression. Int J Mol Sci. 2023;24: 16893.

40. Dorff TB, Narayan V, Forman SJ, Zang PD, Fraietta JA, June CH, et al. Novel Redirected T-Cell Immunotherapies for Advanced Prostate Cancer. Clin Cancer Res. 2022;28: 576–584.

41. Griss J, Viteri G, Sidiropoulos K, Nguyen V, Fabregat A, Hermjakob H. ReactomeGSA - Efficient Multi-Omics Comparative Pathway Analysis. Mol Cell Proteomics. 2020;19: 2115–2125.

42. Jin S, Guerrero-Juarez CF, Zhang L, Chang I, Ramos R, Kuan CH, et al. Inference and analysis of cell-cell communication using CellChat. Nat Commun. 2021;12: 1088.

43. Hong B, Li Y, Yang R, Dai S, Zhan Y, Zhang WB, et al. Single-cell transcriptional profiling reveals heterogeneity and developmental trajectories of Ewing sarcoma. J Cancer Res Clin Oncol. 2022;148: 3267–3280.

44. Nguyen-Nielsen M, Borre M. Diagnostic and Therapeutic Strategies for Prostate Cancer. Semin Nucl Med. 2016;46: 484–490.

45. Tang C, Huang Z, Li H, Zhang R, Yu G, Kong J, et al. EXPAR and Au–Ag mushroom-shaped SERS probe assisted detection of exosomal miR-375 in prostate cancer. Sensors & Diagnostics. 2023;2: 1553–1560.

46. Zhou C, Zhu HL, Duan Y. Targeting Histone Acetyltransferase MOZ/KAT6A as a New Avenue for Hematological Tumor Therapy. Curr Top Med Chem. 2020;20: 333–335.

47. Gryder BE, Akbashev MJ, Rood MK, Raftery ED, Meyers WM, Dillard P, et al. Selectively targeting prostate cancer with antiandrogen equipped histone deacetylase inhibitors. ACS Chem Biol. 2013;8: 2550–2560.

48. Bee A, Brewer D, Beesley C, Dodson A, Forootan S, Dickinson T, et al. siRNA knockdown of ribosomal protein gene RPL19 abrogates the aggressive phenotype of human prostate cancer. PLoS One. 2011;6: e22672.

49. Chen X, Cao Q, Liao R, Wu X, Xun S, Huang J, et al. Loss of ABAT-Mediated GABAergic System Promotes Basal-Like Breast Cancer Progression by Activating Ca(2+)-NFAT1 Axis. Theranostics. 2019;9: 34–47.

50. Lee JV, Berry CT, Kim K, Sen P, Kim T, Carrer A, et al. Acetyl-CoA promotes glioblastoma cell adhesion and migration through Ca(2+)-NFAT signaling. Genes Dev. 2018;32: 497–511.

51. Chen WL, Chang YL, Lin SF, Protzer U, Isogawa M, Yang HC, et al. Differential regulation of calcium-NFAT signaling pathway by Akt isoforms: unraveling effector dynamics and exhaustion of cytotoxic T lymphocytes in tumor microenvironment. J Immunother Cancer. 2025;13: e009827.

52. Neubauer E, Latif M, Krause J, Heumann A, Armbrust M, Luehr C, et al. Up regulation of the steroid hormone synthesis regulator HSD3B2 is linked to early PSA recurrence in prostate cancer. Exp Mol Pathol. 2018;105: 50–56.

53. Li CY, Huang SP, Chen YT, Wu HE, Cheng WC, Huang CY, et al. TNFRSF13B is a potential contributor to prostate cancer. Cancer Cell Int. 2022;22: 180.

54. Guo H, Vuille JA, Wittner BS, Lachtara EM, Hou Y, Lin M, et al. DNA hypomethylation silences anti-tumor immune genes in early prostate cancer and CTCs. Cell. 2023;186: 2765–2782. e2728.

55. Gabrilovich DI, Ostrand-Rosenberg S, Bronte V. Coordinated regulation of myeloid cells by tumours. Nat Rev Immunol. 2012;12: 253–268.

56. Kumar V, Patel S, Tcyganov E, Gabrilovich DI. The Nature of Myeloid-Derived Suppressor Cells in the Tumor Microenvironment. Trends Immunol. 2016;37: 208–220.

57. Di Mitri D, Toso A, Chen JJ, Sarti M, Pinton S, Jost TR, et al. Tumour-infiltrating Gr-1+ myeloid cells antagonize senescence in cancer. Nature. 2014;515: 134–137.

58. Calcinotto A, Spataro C, Zagato E, Di Mitri D, Gil V, Crespo M, et al. IL-23 secreted by myeloid cells drives castration-resistant prostate cancer. Nature. 2018;559: 363–369.

59. Wang G, Lu X, Dey P, Deng P, Wu CC, Jiang S, et al. Targeting YAP-Dependent MDSC Infiltration Impairs Tumor Progression. Cancer Discov. 2016;6: 80–95.

60. Lu X, Horner JW, Paul E, Shang X, Troncoso P, Deng P, et al. Effective combinatorial immunotherapy for castration-resistant prostate cancer. Nature. 2017;543: 728–732.

61. Veglia F, Sanseviero E, Gabrilovich DI. Myeloid-derived suppressor cells in the era of increasing myeloid cell diversity. Nat Rev Immunol. 2021;21: 485–498.

62. Bezzi M, Seitzer N, Ishikawa T, Reschke M, Chen M, Wang G, et al. Diverse genetic-driven immune landscapes dictate tumor progression through distinct mechanisms. Nat Med. 2018;24: 165–175.

63. Zhao D, Cai L, Lu X, Liang X, Li J, Chen P, et al. Chromatin Regulator CHD1 Remodels the Immunosuppressive Tumor Microenvironment in PTEN-Deficient Prostate Cancer. Cancer Discov. 2020;10: 1374–1387.

64. Liu Y, Zhao F, Wang Q, Zhao Q, Hou G, Meng Q. Current Perspectives on Paclitaxel: Focus on Its Production, Delivery and Combination Therapy. Mini Rev Med Chem. 2023;23: 1780–1796.

65. Miller AV, Hicks MA, Nakajima W, Richardson AC, Windle JJ, Harada H. Paclitaxel-induced apoptosis is BAK-dependent, but BAX and BIM-independent in breast tumor. PLoS One. 2013;8: e60685.

66. Tannock IF, de Wit R, Berry WR, Horti J, Pluzanska A, Chi KN, et al. Docetaxel plus prednisone or mitoxantrone plus prednisone for advanced prostate cancer. N Engl J Med. 2004;351: 1502–1512.

67. Ramaswamy B, Puhalla S. Docetaxel: a tubulin-stabilizing agent approved for the management of several solid tumors. Drugs Today (Barc). 2006;42: 265–279.

68. Ruiz de Porras V, Wang XC, Palomero L, Marin-Aguilera M, Solé-Blanch C, Indacochea A, et al. Taxane-induced Attenuation of the CXCR2/BCL-2 Axis Sensitizes Prostate Cancer to Platinum-based Treatment. Eur Urol. 2021;79: 722–733.

69. Varga J, Greten FR. Cell plasticity in epithelial homeostasis and tumorigenesis. Nat Cell Biol. 2017;19: 1133–1141.

70. Tiwari R, Manzar N, Ateeq B. Dynamics of Cellular Plasticity in Prostate Cancer Progression. Front Mol Biosci. 2020;7: 130.

71. Nagl L, Horvath L, Pircher A, Wolf D. Tumor Endothelial Cells (TECs) as Potential Immune Directors of the Tumor Microenvironment - New Findings and Future Perspectives. Front Cell Dev Biol. 2020;8: 766.

72. Hida K, Maishi N, Torii C, Hida Y. Tumor angiogenesis--characteristics of tumor endothelial cells. Int J Clin Oncol. 2016;21: 206–212.

73. Lin BB, Huang Q, Yan B, Liu M, Zhang Z, Lei H, et al. An 18-gene signature of recurrence-associated endothelial cells predicts tumor progression and castration resistance in prostate cancer. Br J Cancer. 2024;131: 870–882.

74. Flammiger A, Bayer F, Cirugeda-Kühnert A, Huland H, Tennstedt P, Simon R, et al. Intratumoral T but not B lymphocytes are related to clinical outcome in prostate cancer. Apmis. 2012;120: 901–908.

75. Palucka AK, Coussens LM. The Basis of Oncoimmunology. Cell. 2016;164: 1233–1247.

76. La Manna MP, Di Liberto D, Lo Pizzo M, Mohammadnezhad L, Shekarkar Azgomi M, Salamone V, et al. The Abundance of Tumor-Infiltrating CD8(+) Tissue Resident Memory T Lymphocytes Correlates with Patient Survival in Glioblastoma. Biomedicines. 2022;10: 2454.

77. Xue C, Zhou Q, Zhang P, Zhang B, Sun Q, Li S, et al. MRI histogram analysis of tumor-infiltrating CD8+ T cell levels in patients with glioblastoma. Neuroimage Clin. 2023;37: 103353.

78. Lesokhin AM, Hohl TM, Kitano S, Cortez C, Hirschhorn-Cymerman D, Avogadri F, et al. Monocytic CCR2(+) myeloid-derived suppressor cells promote immune escape by limiting activated CD8 T-cell infiltration into the tumor microenvironment. Cancer Res. 2012;72: 876–886.

79. Zhang Md J, Zhang Md L, Yang Md Y, Liu Md Q, Ma Md H, Huang Md A, et al. Polymorphonuclear-MDSCs Facilitate Tumor Regrowth After Radiation by Suppressing CD8(+) T Cells. Int J Radiat Oncol Biol Phys. 2021;109: 1533–1546.

80. Di Pilato M, Kim EY, Cadilha BL, Prüßmann JN, Nasrallah MN, Seruggia D, et al. Targeting the CBM complex causes T(reg) cells to prime tumours for immune checkpoint therapy. Nature. 2019;570: 112–116.

81. Ban Y, Mai J, Li X, Mitchell-Flack M, Zhang T, Zhang L, et al. Targeting Autocrine CCL5-CCR5 Axis Reprograms Immunosuppressive Myeloid Cells and Reinvigorates Antitumor Immunity. Cancer Res. 2017;77: 2857–2868.

82. Kang YW, Choi D, Moon D, Lee KJ, Oh Y, Yang J, et al. Enabling immune checkpoint blockade efficacy in T-lymphopenia by restoring CD8 T cell dynamics with IL-7 cytokine therapy. Front Immunol. 2024;15: 1477171.

83. Wang B, Zhang W, Jankovic V, Golubov J, Poon P, Oswald EM, et al. Combination cancer immunotherapy targeting PD-1 and GITR can rescue CD8(+) T cell dysfunction and maintain memory phenotype. Sci Immunol. 2018;3: eaat7061.

84. Nagahisa T, Yamaguchi S, Kosugi S, Homma K, Miyashita K, Irie J, et al. Intestinal Epithelial NAD+ Biosynthesis Regulates GLP-1 Production and Postprandial Glucose Metabolism in Mice. Endocrinology. 2022;163.

85. Wang Q, Liu W, Zhou H, Lai W, Hu C, Dai Y, et al. Tozasertib activates anti-tumor immunity through decreasing regulatory T cells in melanoma. Neoplasia. 2024;48: 100966.

